# LLM-Guided Weak Supervision Improves Active Learning for Abstract Screening

**DOI:** 10.1101/2025.08.24.25334314

**Authors:** Opeoluwa Akinseloyin, Xiaorui Jiang, Vasile Palade

## Abstract

The abstract screening step, determining which studies are relevant for systematic reviews, can require months of expert labour. Active learning has emerged as a promising automation strategy, but its effectiveness is severely limited by the cold-start problem: without labelled training examples at the outset, classifiers initialise poorly and frequently fail to retrieve the 95% of relevant studies required for valid evidence synthesis. This paper introduces an LLM-based weak supervision framework that resolves the cold-start problem by using large language models to score and rank all candidate studies before any human annotation occurs. Documents at the top and bottom of this ranking are designated as pseudo-labelled positives and negatives, providing a fully informed initialisation of the active learning cycle without manual labelling effort. We evaluate the framework through 4,032 simulation experiments across 28 systematic reviews about clinical intervention and diagnostic technology assessment, spanning six query strategies and 14 pseudo-labelling ratios. Pseudo-labelled initialisation raises classifier recall from 0.136 to 0.996 and AUC-ROC from 0.640 to 0.909, confirming that initialisation quality is the primary determinant of screening success. Integrated with active learning, success rates reach 95.8-98.2% and total recall 99.1-99.3%, compared to 72.0% under random initialisation (Cohen’s *d* ≈ 0.86), with near-perfect success achievable across dataset sizes through size-adaptive ratio selection. These results establish LLM-based pseudo-labelling as a robust, strategy-agnostic initialisation prerequisite that reduces manual screening burden while ensuring near-complete evidence retrieval, a capability with broad implications for evidence synthesis across medicine, policy, and the social sciences.

**Highlights:**

- **What is already known.** Active learning, supported by tools such as ASReview, accelerates systematic review screening but suffers from three persistent limitations: cold-start labelling requirements, severe class imbalance, and high performance variability. The SAFE procedure (Boetje and van de Schoot, 2024) addresses stopping but still presumes human-labelled seeds. Neither direct LLM classification nor active learning initialised with random sampling can guarantee adequate recalls of included studies for trustworthy review—near total or at least 95% recall as an accepted real-world standard..
- **What is new.** This is the first systematic investigation of how its performance and reliability in abstract screening are affected by the initialisation method and query strategies. This study reframes LLM-based pseudo-labelling as a universal initialisation mechanism for active learning: LLMs rank the candidate pool, and the top- and bottom-ranked fractions are pseudo-labeled as positives and negatives, removing all upfront annotation. Four novel query strategies are proposed and evaluated: two by repetitive alternation between uncertainty- and certainty-based sampling, and two by one-off switch between uncertainty- and certainty-based sampling. Our experiments show that replacing random initialisation with LLM-based pseudo-labelling makes SAFE highly reliable stopping criteria for active learning-based abstract screening, achieving 95% recall on all reviews using the optimal setup(s). Contrary to prior studies, initialisation quality, not query strategy or convergence speed, is the primary determinant of screening reliability.
- **Potential impact for Research Synthesis Methods readers.** For reviewers, the proposed approach can be used as a pre-screener which safely removes a significant portion of candidate studies (on average approximately 40% over the tested reviews) from manual screening at near-total recall, with no upfront annotation and at LLM API costs of a few cents per review, however, at a sacrifice of lacking explanation and interaction with AI. LLM-based pseudo-laballing significantly improves the SAFE procedure as a set of stopping criteria for active learning and makes it truly safe and efficient in practical use. For methodologists, initialisation, rather than query strategy, deserves first-class attention. For tool developers, pseudo-labelling is a drop-in, classifier- and extractor-agnostic layer compatible with platforms such as ASReview without algorithmic redesign.

## 1. Introduction

A systematic review (SR) is an indispensable tool for synthesising existing evidence on a research topic, particularly in evidence-based medicine where its findings directly inform clinical practice guidelines and health policy (Michelson and Reuter, 2019). Conducting a rigorous SR is, however, extraordinarily resource-intensive. One widely cited estimate places the cost of a single SR at 67.3 weeks of effort and millions of dollars for academic institutions and pharmaceutical companies (Michelson and Reuter, 2019). A cost-effectiveness analysis further estimated that each reviewer requires between 83 and 125 hours to screen 5,000 references, at a cost of approximately $17,000 (at 2013 prices) (Shemilt et al., 2016). With best-practice guidelines recommending no more than three hours of screening per reviewer per day to maintain quality (Polanin et al., 2019), even abstract screening alone can consume over a month of dedicated effort. The global volume of published research is growing at approximately 4% annually, with over 2.5 million new articles indexed each year (Bastian et al., 2010), making the manual synthesis of evidence increasingly unsustainable.

Responding to this challenge, substantial effort has been devoted to AI-aided systematic review tools that leverage machine learning to automate or semi-automate the abstract screening step (van Dinter et al., 2021b; Dos Santos et al., 2023; Ofori-Boateng et al., 2024a). However, several interconnected grand challenges have prevented these tools from achieving reliable, widespread deployment.

● **Challenge 1:** Lack of labelled data. The success of machine learning hinges on the availability of high-quality labelled training data, but each SR constitutes a distinct dataset with no annotations at the outset. Acquiring such labels is resource-intensive, particularly when expert clinical knowledge is required (Garg and Kalai, 2018).
● **Challenge 2:** Non-generalisability across review topics. Models trained on one SR are rarely transferable to another because each review addresses a different research question, study design, and outcome of interest. This necessitates manual annotation for each new review, undermining the efficiency gains automation is intended to provide.
● **Challenge 3:** Extreme class imbalance. Most of the time, abstract screening is inherently a highly imbalanced classification task; the ratio of relevant to irrelevant studies can be as high as 1: several hundred or over a thousand (Szucs and Papp, 2022). This makes it exceptionally difficult to discover positive examples (relevant or included studies) with minimal supervision and to build a well-calibrated classifier.
● **Challenge 4:** Non-negotiable recall requirement. A systematic review that misses relevant studies produces biased, potentially invalid evidence synthesis. International guidelines mandate that automated tools should achieve near total recall (e.g., 95%) of relevant studies (Kanoulas et al., 2019). This requirement is far more stringent than is typical in general text classification tasks.
● **Challenge 5:** The cold-start and stopping problem. Even when active learning partially addresses the annotation bottleneck, its practical deployment faces two compounding difficulties. First, the cold-start problem, having no labelled data to initialise the classifier at the beginning of a review, forces random or heuristic initialisation strategies that perform poorly under class imbalance. Second, reliably determining when to stop the active learning cycle without missing relevant records requires robust stopping criteria that must be calibrated for the systematic review context, but this has proved extremely difficult(Boetje and van de Schoot, 2024; Callaghan et al., 2024).

Active learning has become the predominant strategy to address these challenges by iteratively selecting the most informative examples for manual labelling, thereby minimising the total annotation burden (Lewis, 1995; Wallace et al., 2010b; van de Schoot et al., 2021). ASReview, the high-profile and most widely adopted active learning-based open-source abstract screening tool, has gained significant attention since its introduction in 2021 and underpinned numerous simulation benchmarks and methodological evaluations (van de Schoot et al., 2021; Ferdinands et al., 2023; Teijema et al., 2025). Despite this progress, large-scale simulation studies consistently reveal that active learning performance is highly variable: under random initialisation, the fraction of reviews that achieve 95% recall varies widely across datasets, and a substantial proportion fail to do so (Teijema et al., 2025; Ferdinands et al., 2023). Structured stopping criteria, such as the SAFE (Systematic Active learning Framework for Evidence synthesis) procedure (Boetje and van de Schoot, 2024), have been proposed to make the endpoint more reliable. Still, it has neither addressed the upstream initialisation problem that causes many reviews to underperform before the stopping criterion is ever reached, nor conducted a large-scale evaluation of SAFE’s reliability, particularly in achieving near-total recall for SRs.

This paper fills these gaps by introducing a zero-annotation initialisation framework for active learning based on LLM-generated pseudo-labels. The proposed approach employs LLMs to score and rank all candidate studies according to each review’s selection criteria before the active learning cycle begins. The top T% of ranked documents are pseudo-labelled as positives and the bottom B% as negatives, producing a well-balanced, high-coverage initial training set without any human annotation. This transforms the cold-start problem from a fundamental obstacle into a solved initialisation step.

Crucially, we evaluate the approach not as a competing active learning method but as a universal initialisation layer. Experiments are conducted using a modified version of the ASReview open-source Python library as the simulation platform, with the SAFE stopping criterion standardised across all conditions, so that only the initialisation strategy varies. This design demonstrates that pseudo-labelling enhances any active learning configuration, whether it uses certainty-based sampling, uncertainty-based sampling, or a hybrid strategy alternating between the two, without modifying any aspect of the downstream active learning algorithm.

In summary, this study has the following main contributions:

1. We propose a novel LLM-based pseudo-labelling framework for zero-annotation initialisation of active learning in systematic review abstract screening, which eliminates the need for upfront manual annotation.
2. We demonstrate that pseudo-labelling is a universal initialisation prerequisite: it improves the reliability and efficiency of every active learning strategy evaluated, including both certainty- and uncertainty-based sampling.
3. We conduct a large-scale simulation study of initialisation quality in systematic review automation to date, spanning 4032 experiments across 28 datasets, 6 query strategies, and 14 pseudo-labelling ratios.
4. We demonstrate that integrating LLM-based pseudo-labelling into SAFE transforms it into a highly reliable and practically deployable screening framework. As part of this integration, we provide a direct empirical comparison of the End-of-Stage-S classifier (LLM-only, zero human annotation) versus the End-of-Stage-A classifier (SAFE-guided active learning) performance, revealing that LLM quality, not active learning algorithm design, is the primary bottleneck in the combined pipeline.

## 2. Related Work

### 2.1. Machine Learning for Abstract Screening

There have been nearly two decades of research in using machine learning to automate or semi-automate systematic review screening. In the seminal work of Cohen et al. (2006), abstract words, MeSH terms, and MEDLINE publication tags were used to construct document representations, and a voting perceptron was trained to predict inclusion/exclusion. Wallace et al. (2010b) extended this with bag-of-biomedical-words features based on UMLS terms and trained a support vector machine (SVM). A 2021 analysis of 41 studies confirmed that SVM and Naive Bayes were the most popular classifiers and TF-IDF the most common feature extraction method (van Dinter et al., 2021b). More recent comprehensive reviews covering natural language processing, machine learning, and deep learning approaches confirm that the field has grown substantially, but no single method reliably achieves high recall across diverse review types (Ofori-Boateng et al., 2024a; Dos Santos et al., 2023). The most prominent challenge across all approaches remains the unavailability of labelled data at the start of a review, which our work directly addresses.

### 2.2. Active Learning for Systematic Review Screening

Active learning is the dominant strategy for reducing the annotation burden in systematic review screening, and has been formalised as the core mechanism in tools including ASReview (van de Schoot et al., 2021), Abstrackr (Wallace et al., 2012), RobotAnalyst (Przybyla et al., 2018), and SWIFT-Active Screener (Howard et al., 2020). The key design choices in active learning are the query strategy, which determines which unlabelled documents to present to the reviewer, and the construction of the initial training set.

Query strategies span a spectrum between certainty-based sampling (presenting documents that the classifier is most confident are relevant) and uncertainty-based sampling (presenting documents near the classifier’s decision boundary). Certainty-based strategies tend to find relevant documents faster initially but can converge prematurely; uncertainty-based strategies explore more broadly but may take longer to achieve high recall (Miwa et al., 2014; Howard et al., 2020; Ofori-Boateng et al., 2024b). Hybrid strategies that switch between modes at different stages of the screening cycle have been proposed to capture the benefits of both (Cormack and Grossman, 2016). A comprehensive simulation benchmark across 28 datasets confirmed that these differences in strategies are real and significant, but that no single strategy dominates across all review types (Teijema et al., 2025).

The success of active learning, however, is critically determined by the quality of the initial training dataset (Hwang et al., 2024; Chen et al., 2023). Diversity-based approaches apply clustering algorithms to select representative instances for labelling (Kang et al., 2004; Zhu et al., 2008), while border-based methods select samples near cluster boundaries as potentially more discriminative (Yuan et al., 2011). A hybrid of centre-based and border-based selection was shown to outperform individual strategies (Yuan et al., 2011). These methods all suffer from class imbalance in the context of systematic reviews. Without pre-existing knowledge of which documents are relevant, discovering a representative set of positive examples through random or cluster-based selection is unreliable. Our approach resolves this by using LLMs to rank documents before the active learning cycle begins, ensuring a balanced and high-coverage initial training set.

### 2.3. The Cold-Start Problem in Systematic Review Active Learning

The cold-start problem in active learning refers to the difficulty of constructing an effective initial training set when no labelled data is available. It is a well-recognised challenge in the broader active learning literature (Souza et al., 2017; Szucs and Papp, 2022), but is particularly acute in systematic review screening due to the extreme class imbalance. Simulation studies using ASReview consistently show that initialisation with a single relevant and irrelevant record, the minimal feasible starting point, produces high variance in performance across datasets: some reviews converge quickly, others fail to find relevant records for dozens of iterations before the classifier learns the decision boundary (Ferdinands et al., 2023; Teijema et al., 2025; Byrne et al., 2024).

Recent work has attempted to address cold-start directly. Chen et al. (2023) examined the cold-start problem in medical active learning using self-supervised pre-training, demonstrating that better initialisation improves downstream performance. Hwang et al. (2024) proposed leveraging self-supervised language model representations to more effectively construct initial datasets. Szucs and Papp (2022) developed a zero-initialised unsupervised active learning method using entropy-based sampling for imbalanced problems. Our approach extends this line of work to the systematic review domain, using LLMs’ semantic understanding of inclusion criteria, rather than generic clustering, to generate high-quality pseudo-labels that simultaneously address both the cold-start problem and class imbalance.

### 2.4. Stopping Criteria and the SAFE Procedure

A complementary but equally important challenge in active learning for systematic reviews is determining when to stop screening. Without reliable stopping criteria, reviewers face an inherent tension: stopping too early risks missing relevant records, while continuing unnecessarily increases workload. This challenge has received increasing attention as active learning tools have moved from research environments to operational deployment (Gates et al., 2020; Callaghan et al., 2024; Pilz et al., 2024). For example, Callaghan et al. (2024) provide a broader evaluation of stopping criteria for computer-assisted screening, confirming that the choice of stopping criterion significantly affects both recall and screening burden and emphasising the need for criteria that are robust across diverse datasets.

Recently, Boetje and van de Schoot (2024) introduced the SAFE (Systematic Active Learning Framework for Evidence Synthesis) procedure, a structured, four-stage stopping protocol designed for the practical deployment of active learning in systematic reviews. Stage S initialises the classifier using a small random sample of the dataset (approximately 1% as suggested by Boetje and van de Schoot). Stage A applies active learning with a compound four-criterion stopping rule: (i) all pre-specified key papers have been identified, (ii) at least twice the expected number of relevant records have been screened, (iii) a minimum of 10% of the total dataset has been reviewed, and (iv) no relevant records have been found in the last 50 consecutive screenings. Stage F applies a more advanced model on the Stage S and Stage A labels to continue screening until the Stage F stopping criterion is met. Stage E evaluates overall quality. The SAFE procedure has been validated as a practical, conservative, and reproducible stopping rule that balances recall and efficiency across a range of systematic review types (Boetje and van de Schoot, 2024).

Subsequently the SAFE procedure has been adopted in real-world reviews as a truly “safe” method despite the inadequacy of convincing large-scale empirical results. For example, Sagot-Better (2026) used SAFE and claimed that it allowed the author “to determine when to stop the active learning-aided screening while adhering to the PRISMA guidelines”. Several follow-up studies evaluated the SAFE procedure. While the purpose of König et al. (2024) were to derive recommendations for three non-SAFE stopping rules tailored to different prevalence ratios of positive and negative samples, they proposed a seven-step AI-aided screening pipeline where the sixth and seventh steps were essentially borrowed from SAFE’s last two stages. Swab (2026) directly evaluated the performance of SAFE through a retrospective study using the ASReview software and its default setup (TFIDF + Naive Bayes; the same setup as used by the current paper), but failed on 4 out of 10 reviews (<95% recall) and severely underperformed on three (<90%). Verdonschot et al. (2026) performed a more comprehensive performance evaluation on four large pharmacy reviews by varying choices of feature extractor, classifier model, prior knowledge, and the label set (screening decisions based on title-abstract vs fulltext). SAFE failed on one of the four reviews with the recommended best parameter combination in ASReview Lab v2.0 and failed on two with the default setup, when full-text screening labels were used for these retrospective studies. Different from Verdonschot et al’s, our work focuses on the initialisation method and integrates the SAFE Stage A stopping criteria as the standardised endpoint across all experimental conditions, ensuring that differences in observed performance are attributable to initialisation quality rather than to variable stopping decisions.

### 2.5. Simulation Benchmarks and Large-Scale Evaluation

Rigorous evaluation of active learning tools for systematic reviews requires simulation studies capable of replicating the human screening process at scale. Ferdinands et al. (2023) conducted a simulation study across six datasets comparing four classifiers (Naive Bayes, logistic regression, SVM, random forest) and two feature extraction methods (TF-IDF, Doc2Vec), introducing the Average Time to Discovery (ATD) metric to capture performance across the entire active learning cycle rather than at an arbitrary cutoff. Teijema et al. (2025) expanded this significantly, evaluating 325 classifier-feature extractor combinations across multiple datasets and reporting substantial variability both across models and across datasets, concluding that no single configuration universally dominates.

The CLEF eHealth 2019 Task 2 benchmark (Kanoulas et al., 2019) has become the most widely used dataset for evaluating active learning tools in empirical medicine, providing 28 systematic reviews with ground-truth inclusion/exclusion labels across Intervention and Diagnostic Test Accuracy (DTA) review types. This benchmark underpins the simulation studies of both Teijema et al. (2025) and the present work. Our experimental design extends this benchmark by systematically varying initialisation strategy, pseudo-labelling ratio, and query strategy across all 28 datasets, enabling the most comprehensive evaluation of initialisation quality in the systematic review automation literature to date.

### 2.6. Large Language Models for Abstract Screening

Large language models (LLMs) have attracted substantial recent interest as direct tools for abstract screening. However, their reliability as standalone classifiers has been inconsistent. Guo et al. (2024) applied GPT -4 using engineered inclusion/exclusion-criterion prompts, but the average sensitivity was only 76%. In Oami et al. (2024), the pooled sensitivity across five reviews was only 49%. Combining multiple LLMs’ decisions through ensemble methods partially mitigates individual-model bias and improves recall (Li et al., 2024; Sanghera et al., 2025), but at a substantially increased computational cost.

A more promising direction treats LLMs as ranking rather than classifying tools. Dennstadt et al. (2024) and Issaiy et al. (2024) demonstrated that LLM-based ranking on a Likert scale can achieve more reliable prioritisation than binary classification. Wang et al. (2024) proposed a zero-shot generative approach using LLMs to automate screening. Akinseloyin et al. (2024) developed a question-answering framework in which each inclusion criterion is transformed into an explicit question and answered by the LLM, demonstrating that criterion-level reasoning significantly improves reliability compared with single-prompt classification. Our approach builds on this question-answering framework (Akinseloyin et al., 2024) to generate document relevance scores. Still, it uses these scores to generate pseudo-labels rather than for direct classification, thereby leveraging the LLM’s semantic understanding while bypassing its reliability limitations as a primary decision-maker.

## 3. Methodology

This section describes the LLM-based pseudo-labelling framework, its integration into the SAFE active learning procedure, the experimental simulation design, and the evaluation metrics and statistical framework employed throughout the study.

### 3.1. LLM-Based Screening Prioritisation

#### 3.1.1. Problem Definition

For an SR, suppose the unlabelled document set, i.e., the candidate studies (here titles and abstracts) for screening, is denoted by 𝒟 = {*d*_1_, *d*_2_, ⋯, *d_N_*}, where *d_i_* is the *i*-th document, and 𝑁 can be quite large, from thousands to tens of thousands. The abstract screening task aims to train a machine learning model M that assigns a binary label to each document: ℳ: 𝒟 ⟼ 𝒴, where 𝒴 = {0,1}, with “0” indicating exclusion and “1” indicating inclusion. To start the active learning cycle for abstract screening, we must obtain a small set of initial labels as the training set and split the dataset into two parts: ℒ = {(*d*_1_, 𝑦_1_), (*d*_2_, 𝑦_2_), ⋯, (*d_l_*, 𝑦*_l_*)} is the set of labeled samples, where 𝑦_*j*_ ∈ {0, 1} (∀𝑗 ∈ {1, ⋯, 𝑙}), and 𝒰 = {*d*_*1*+1_, ⋯, *d_N_*} is the set of unlabeled samples. It is challenging to obtain a high-quality labelled set𝐿to train a good classifier to start the active learning cycle, and the performance of the initial classifier significantly impacts active learning’s effectiveness in real-world screening practice, the latter of which will be empirically demonstrated in the Results section.

#### 3.1.2. Scoring Using Large Language Models

Each SR specifies several selection criteria that every included study must satisfy. Following Akinseloyin et al. (2024), each criterion is transformed into a question, again automatically by LLMs, with three possible answers: “Yes”, “No” or “N/A” (not answerable, unsure or neutral). Let 𝒬 = {𝑞_1_, ⋯, 𝑞*_K_*} denote the set of inclusion criteria questions. Based on the questions, an LLM is used to score each study. For each document *d_i_*, an LLM is first used to produce the answers to all the questions: 𝒜_*i*_ = =𝑎*_i,i_*, ⋯, 𝑎*_K,i_*. Where 𝑎*_k,i_* is the answer for the *k*-th question 𝑞_*k*_ on the *i*-th document *d*_*i*_. A “Yes” answer starts with the answer word “Positive” followed by an LLM-generated reason for the answer, while the “No” and “N/A” answers start with “Negative” and “Neutral” respectively. Then, an initial score is assigned to measure the adherence of document *d_i_* to the *k*-th criterion, denoted by 𝑠(*d_i_*, 𝑞*_k_*), based on its answer 𝑎*_k,i_* and the reasoning text for question 𝑞*_k_*. “Yes” and “No” answers will result in scores near 1 and 0, respectively, while a “N/A” answer will result in a score near 0.5. See Supplementary File 1 Section A for details of the prompt and an example. In this paper, the initial score is the probability that an answer text has positive sentiment, as estimated by a BART-based sentiment analyser (Muthukumar, 2021).

Using LLM-generated answers alone does not produce the optimal results (Akinseloyin et al., 2024; Guo et al., 2024; Oami et al., 2024), so the following hypothesis was engineered to re-rank LLM-based results: Given an included study, there must be a certain part of it that matches the corresponding inclusion criterion, so it is natural to assume a high semantic relevance between the study and the criterion question. Therefore,

the re-ranked score is calculated by averaging the initial score with the semantic relevance between the study and the corresponding question, denoted by 𝑠̅(*d_i_*, 𝑞*_k_*):

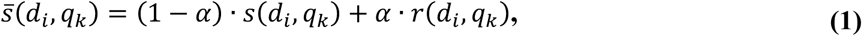

Where 𝑟(*d_i_*, 𝑞*_k_*)is the semantic relevance between the *i-*th document and the *k*-th criterion question and 𝛼 ∈ (0, 1) is a controlling parameter. In this paper, 𝑟(*d_i_*, 𝑞*_k_*) is approximated by the cosine similarity between the representation vectors (i.e., text embeddings) of document *d_i_* and criterion question 𝑞*_k_*. The initial score of each document is defined as the average of its scores with respect to all questions that represent the complete inclusion criteria:

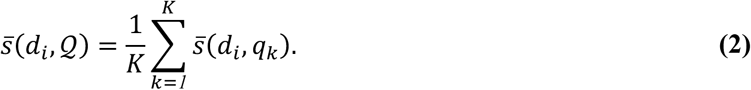

Because an included study is expected to meet all selection criteria, we assume a high semantic relevance between it and the inclusion-criteria paragraph in the SR protocol, denoted by 𝒬. So, the document score can be further adjusted using this document-level semantic relevance (denoted by) as follows:

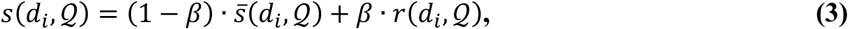

Where 𝑟(*d_i_*, 𝑖) is the cosine similarity between the text embeddings of the document and the selection criteria paragraph, and 𝛽 ∈ (0, 1) is another controlling parameter. In our experiments, we simply used 𝛼 = 𝛽 = 0.5.

#### 3.1.3. Scoring Using Large Language Models

Following Akinseloyin et al. (2026), an ensemble of LLMs, denoted as 𝒢 = {𝑔*_1_*, ⋯, 𝑔*_M_*} with each 𝑔*_j_* (𝑗 ∈ {1, 2, ⋯, 𝑀}) being an LLM, was employed to reduce the potential bias of the individual LLMs. For each SR with the selection-criteria questions (i.e., selection-criteria paragraph) 𝒬 and each candidate study di, the ensemble averages the scores assigned by each individual LLM *g*, denoted as 𝑠*_g_*(*d_i_*, 𝒬), as follows. This method has been proven extremely effective for candidate study prioritisation (Akinseloyin et al., 2026) while maintaining minimal additional costs (Also see Supplementary File 1 Section E).

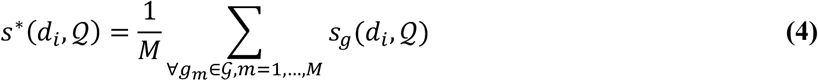

### 3.2. Weakly Supervised Active Learning

#### 3.2.1. Handle Data Imbalance by Pseudo-Labelling

Systematic reviews often exhibit a high degree of class imbalance, where the relevant documents (𝑦 = 1) are significantly outnumbered by the irrelevant ones (𝑦 = *0*). Existing methods struggle to initialise a classifier effectively under such high imbalance. In this paper, we propose generating and using pseudo-labels based on LLM-based document scores obtained using the method described in the prior sections. Suppose we sort the documents in descending order of their scores, forming a ranked list

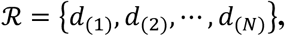

where *d*_(*i*)_ represents the top *i*-th document according to LLM-assigned scores such that ∀*i* ≤ 𝑗, we have 𝑠(*d*_(*i*)_, 𝑄) ≥ 𝑠(*d*_(*j*)_, 𝑄) . The ranking procedure prioritises the documents most likely to be relevant, significantly increasing the chance of identifying positive samples at the top of the ranked list ℛ. We propose to select the top 𝑇% of the ranked documents as (pseudo-labelled) positive samples, denoted as 𝒫, and correspondingly the bottom 𝐵% as (pseudo-labelled) negative samples, denoted as 𝒩. More formally, they are created as follows:

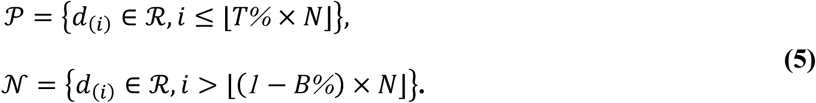

Using the pseudo-labels, we are able to initialise the training set as follows in order to train an initial classifier and initialise the active learning cycle:

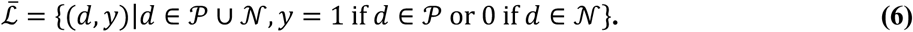

Because the initial training set is built using pseudo-labels, the complete dataset 𝐷 is not annotated with human-assigned labels.

#### 3.2.2. Initialising and Running Active Learning Cycle

After obtaining the pseudo-labelled training set, an initial probabilistic classifier 𝑓 is trained on ℒ. Then batch-based active learning starts: use the classifier to select a batch of the most informative unlabeled samples, send them for human annotation, rebuild the training set with human-assigned labels, and repeat the process iteratively until a predefined stopping condition is met. The informativeness score 𝑠(*d*) assigned to an unlabelled document *d* depends on the query strategy in use. In this study, multiple query strategies are considered. For example, uncertainty-based sampling, which prioritises documents about which the classifier is least confident, and certainty-based sampling, which prioritises documents that the classifier is most confident belong to the positive (relevant) class. The score is defined as follows:

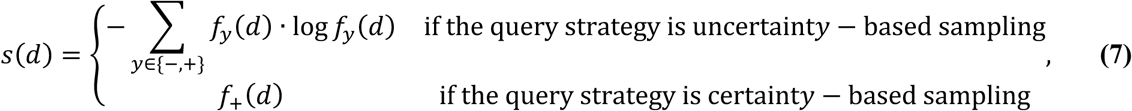

where 𝑓*_y_*(*d*) denotes the posterior probability assigned by classifier 𝑓 to class label 𝑦 (either “–“ or “+”) for document *d*, and 𝑓_+_(*d*) is the posterior probability of the positive (relevant) class. The uncertainty score corresponds to the Shannon entropy of the predicted class distribution (Maximum Entropy sampling), which is maximised when the classifier is most undecided. The certainty score is simply the predicted relevance probability, which is maximised when the classifier is most confident that a document is relevant.

A batch ℬ of *k* documents is then selected from the unlabelled pool 𝒰 by taking the *k* highest-scoring documents:

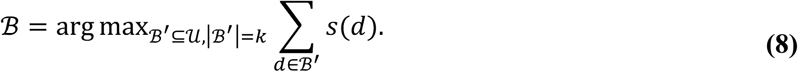

Algorithm 1 summarises how our framework initialises and runs the active learning cycle, called *weakly supervised active learning*, for abstract screening leveraging pseudo-labelling. The overall process is similar to traditional active learning with three notable differences. Firstly, Algorithm 1 uses LLM-assigned pseudo-labels to initialise the training set for the active learning cycle (Step 3). Secondly, during the active learning cycle, the initial pseudo-labelled training set will be improved by new samples, so ℒ^(0)^ is initialised by ℒ̅ (Step 4) and will be iteratively updated. Thirdly, after the active learning cycle is started, the newly sampled documents are labeled and used to improve the training set for re-training: (i) if a sample is not in the pseudo-labelled training set, then simply add it to the training set; (ii) otherwise, replace the label of the original sample with the label assigned by human annotator (Step 11). Note that in Step 11, each selected sample (*d*, 𝑦) will be visited only once, as it will be removed from the unlabelled set. A noteworthy feature of our weakly supervised active learning framework is that it prevents class imbalance from regaining dominance, as the initial set of pseudo-labelled samples (i.e., ℒ̅) is always included in the training data. In addition, our framework enables active learning to select more positive samples throughout the active learning cycle and converge much faster than traditional methods, which is a second factor that alleviates class imbalance.

**Algorithm 1.**
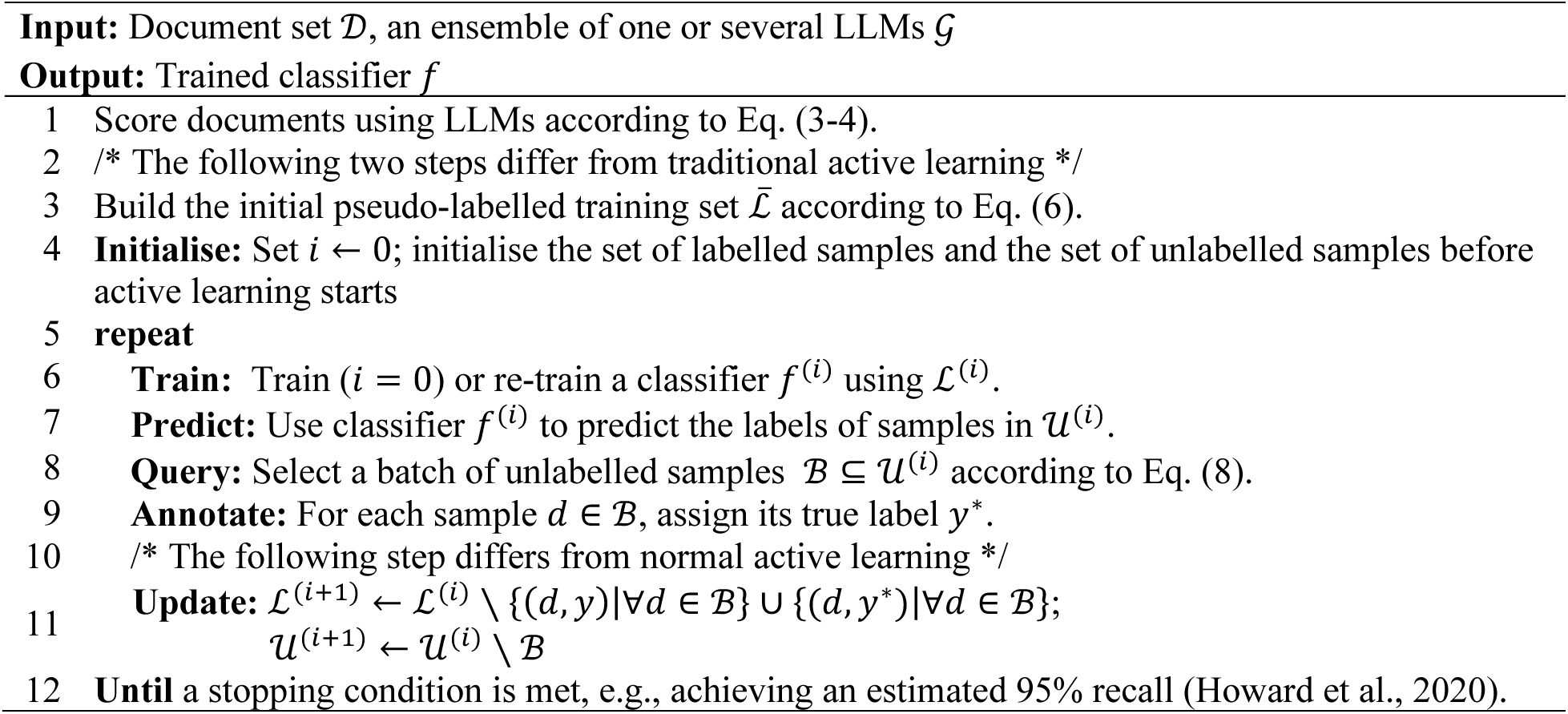
Active Learning Framework for Abstract Screening Based on Pseudo-Labelling.

## 4. Experimental Setup

### 4.1. LLM Setup

For LLM-based scoring and ranking, we used three lightweight and cost-effective LLMs by some of the most famous companies in the LLM industry: GPT-4o mini by OpenAI (more precisely, **gpt-4o-mini-2024-07-18**), Gemini 1.5 Flash by Google AI (more precisely, **gemini-1.5 Flash-001**), and Claude 3 Haiku by Anthropic (more precisely, **claude-3 Haiku-20240307**). The APIs of all three LLMs allow setting the temperature to 0 to ensure replicability, so a temperature of zero was employed throughout our experiments to maintain the stability of the generated responses. For other LLM options, the default values were used. In our experiments, both 𝛼 and 𝛽 are set to 0.5 when calculating the re-ranked document scores (see Equations (1) and (3)).

### 4.2. Datasets

We use 28 datasets from the CLEF eHealth 2019 Task 2, a well-established benchmark for technology-assisted review in empirical medicine (Kanoulas et al., 2019). 20 SRs about clinical intervention trials (Intervention) and 8 SRs about diagnostic test accuracy (DTA) are used for evaluation, with each SR constituting a distinct dataset. Each SR contains a pool of candidate studies (titles and abstracts), and the ground-truth includes/excludes labels. Dataset statistics can be found in Kanoulas et al. (2019). The sizes and protocols for the 28 SRs are available in the supplementary materials.

### 4.3. ASReview Source Code Package for Implementing the Simulation Studies

All active learning experiments are conducted using a modified version of the ASReview open-source Python library (van de Schoot et al., 2021), the most widely adopted open-source active learning tool for systematic reviews, as the simulation platform. ASReview provides a standardised environment for simulating the human reviewer screening process: in each iteration, the model ranks all unlabeled documents by predicted relevance, the top-ranked document is “screened” (its true label revealed), and the model is retrained on the expanded labelled set. This process continues until the stopping criterion is met. Using ASReview grounds our simulation in a tool that practitioners already deploy in real-world systematic reviews, ensuring that our findings speak directly to conditions encountered in practice rather than to an idealised experimental setup. Furthermore, ASReview provides a well-validated, open-source simulation platform with documented default configurations, enabling full reproducibility of our experimental pipeline and transparent evaluation of pseudo-labelling as an initialisation strategy within a standardised framework. We deliberately retain the platform’s default pairing of Naive Bayes with TF-IDF rather than adopting a stronger classifier or denser feature representation. The purpose of this study is to demonstrate the power of pseudo-labelling as a framework, and using a deliberately weak classifier and feature representation isolates the gains attributable to pseudo-labelling itself from those that would otherwise accrue from a more capable model. Any improvements observed under this configuration can therefore be attributed to the initialisation strategy rather than to representational or modelling capacity, with stronger classifier and feature combinations reserved for future works.

Our pseudo-labelling framework integrates into ASReview at the initialisation step. Under the random initialisation baseline, we replicated the default ASReview setup, which performs random sampling ten times and averages the results. With one randomly selected relevant and one irrelevant document (the default ASReview initialisation). Under the pseudo-labelling condition, the initial training set, containing the top *T*% and bottom *T*% of LLM-ranked documents with pseudo-labels, replaces the random initialisation, and active learning then proceeds normally from that starting point. Critically, pseudo-labelled records that are subsequently queried by the active learning algorithm are re-labelled with their true gold-standard labels, ensuring that pseudo-label errors are progressively corrected as the review proceeds.

### 4.4. Integration into the SAFE Procedure

To ensure fair comparison across all initialisation conditions and query strategies, the SAFE Stage A stopping criterion (Boetje and van de Schoot, 2024) is applied as the standardised endpoint for all active learning experiments. Screening stops when the following four-criterion compound rule is simultaneously satisfied: (1) all pre-specified key papers have been identified; (2) at least twice the estimated number of relevant records (estimated from the initial screening fraction) have been screened; (3) at least 10% of the total dataset has been manually reviewed; and (4) no relevant records have been identified in the last 50 consecutive screenings. This stopping rule is conservative by design, intended to minimise the risk of early termination, and its consistent application across all conditions ensures that observed differences in performance reflect initialisation quality rather than variable stopping decisions. However, in our experimental environment, it is impossible to implement Criteria 1 and 2 in their standard form. Criterion 1 cannot be applied because it presupposes that a reviewer has already read through the dataset to designate key papers before screening, which contradicts the cold-start scenario this study addresses. Criterion 2 is equally infeasible, as this criterion relies on an estimate of the total number of relevant records derived from the initially annotated training set. In random initialisation, human reviewers annotate the seed records, providing a verified count of relevant documents from which this estimate can be reasonably drawn. With pseudo-labelling, however, the initial labels are generated by an LLM rather than verified by a human annotator, so the count of relevant records in the training set cannot be confidently ascertained, rendering the estimate unreliable as a stopping threshold. Consequently, our implementation retains only Criteria 3 and 4. This reduced stopping rule remains conservative and is applied consistently across all experimental conditions, ensuring that observed performance differences are attributable to initialisation quality rather than variable stopping behaviour.

Our experimental design deliberately evaluates only Stages S and A of the SAFE procedure. Stage S refers to the LLM-based pseudo-labelling and initial classifier construction (zero human annotation); Stage A refers to the active learning cycle with SAFE stopping. We stop at Stage A for two substantive reasons. First, our primary research question concerns initialisation quality: demonstrating that Stage S and Stage A, when staged together, produce strong, consistent results provides sufficient evidence of the framework’s value without requiring Stage F. Second, Stage F of the SAFE procedure applies a more advanced feature extractor (e.g., sentence-BERT with Random Forests) as a second screening pass and is designed to recover residual relevant records missed in Stage A; since our Stage S and Stage A already achieves near-perfect recalls on nearly all tested reviews across all experimented active learning setups and near-perfect recalls across all tested reviews with the optimised active learning setups, the marginal gain from Stage F would be expected to be minimal.

### 4.5. Classifier and Feature Extraction

Within the active learning cycle (Stage A), we use a Naive Bayes classifier with TF-IDF feature extraction as the base model for all experiments. This corresponds to the default ASReview configuration and is the most widely benchmarked model combination in the systematic review simulation literature (Ferdinands et al., 2023; Teijema et al., 2025; van de Schoot et al., 2021). Class imbalance within the active learning training set is mitigated by dynamic resampling (the “double” balancing strategy in ASReview), which oversamples the positive class to maintain a 1:1 ratio in each training iteration. The use of a standard, well-understood classifier ensures that observed differences between initialisation conditions are attributable to pseudo-labelling quality rather than classifier-specific interactions.

### 4.6. Query Strategies

Six query strategies are evaluated, spanning the full spectrum of approaches used in systematic review active learning tools:

● Pure Uncertainty (PU): Selects the document closest to the classifier’s decision boundary at each iteration. This is the most exploratory strategy, prioritising samples that are most difficult to classify. It is the default in many uncertainty-based tools, such as ASReview’s uncertainty mode.
● Pure Certainty (PC): Selects the document the classifier is most confident is relevant (highest positive posterior probability). This is the default in ASReview’s standard mode and in SWIFT-Active Screener (Howard et al., 2020).

We also design and evaluate four hybrid strategies that blend certainty- and uncertainty-based sampling within the same active learning pipeline. These strategies are introduced to examine whether adaptively transitioning between exploitation and exploration during screening can yield additional gains when pseudo-labelled data is used for initialisation:

● Alternating C→U: Alternates between certainty- and uncertainty-based sampling on consecutive iterations, starting with certainty.
● Alternating U→C: Alternates starting with uncertainty.
● Phase Switch C→U: Uses certainty-based sampling until a fixed fraction of the corpus has been screened, then switches permanently to uncertainty-based sampling.
● Phase Switch U→C: Uses uncertainty-based sampling initially, then switches to certainty-based sampling.

These six strategies collectively represent the principal sampling philosophies in the active learning literature: exploitation (certainty), exploration (uncertainty), and hybrid approaches. Their inclusion enables a comprehensive test of the universality of the pseudo-labelling benefit across diverse algorithmic configurations.

### 4.7. Pseudo-Labeling Ratios

Fourteen pseudo-labelling ratios are evaluated: 2.5%, 5.0%, 7.5%, 10.0%, 12.5%, 15.0%, 17.5%, 20.0%, 22.5%, 25.0%, 27.5%, 30%, 32.5%, and 35%. Each ratio n% corresponds to T = B, so a 12.5% ratio means the top 12.5% of ranked documents are pseudo-labelled as positive and the bottom 12.5% as negative. This range spans from minimal pseudo-labelling (which preserves nearly all records for active learning exploration) to aggressive pseudo-labelling (which provides a large but potentially noisier initial training set). The systematic evaluation of this range allows identification of the optimal ratio and characterisation of the trade-off between pseudo-label quantity and precision.

### 4.8. Evaluation Metrics

#### 4.8.1. Primary Outcomes

The primary evaluation metric is the success rate, defined as the proportion of experiments in which the active learning cycle achieves at least 95% total recall of relevant studies by the time the SAFE stopping criteria are met. This binary threshold succeeds or fails, directly capturing the reliability requirement of systematic review automation: a review that misses more than 5% of relevant studies produces invalid evidence synthesis.

Work Saved over Sampling (WSS) quantifies the reduction in manual screening effort relative to random screening at the same recall level. WSS@Stage A is the actual WSS achieved at the point where the SAFE stopping criterion is met, computed as: WSS = 1 − (*TN* + *FN*)/*N* − *R*%, where *TN* and *FN* are true and false negatives, *N* is the total corpus size, and *R*% is the achieved recall. The actual WSS differs from the theoretical WSS@95% (WSS@95% Theoretical in subsequent sections), which assumes perfect knowledge of the stopping point, and the latter is the more practically meaningful metric because it reflects the real screening burden experienced by the reviewer.

#### 4.8.2. Secondary Outcomes

End-of-Stage-S classifier quality is assessed via recall, precision, F1-score, and AUC-ROC of the initial classifier trained on the pseudo-labelled training set, before any human annotation. Convergence speed is measured as the percentage of the corpus that must be screened to reach 95% and 100% recall under the active learning cycle. Total recall is the final proportion of relevant studies identified at the SAFE stopping point. Screening burden is the proportion of the corpus manually reviewed by the stopping criterion. These secondary outcomes provide a mechanistic understanding of how pseudo-labelling achieves its primary benefits.

### 4.9. Research Questions and Experimental Design

The study is organised around four research questions:

● RQ1: Is LLM-guided pseudo-labelling a reliable and effective approach for abstract (pre-) screening?
● RQ2: What is the optimal configuration for pseudo-labelling towards reliable abstract screening?
● RQ3: Can LLM-based pseudo-labelling enhance the SAFE procedure to achieve reliable and more effective abstract screening?
● RQ4: Does pseudo-labelling allow active learning to converge faster?

The experimental design is fully crossed: 28 datasets × 6 query strategies × 14 pseudo-labelling ratios × 2 initialisation conditions (random and pseudo-labelled) = 4032 experiments. All conditions use the same ASReview simulation platform and the SAFE Stage A-stopping criterion. Statistical comparisons employ Friedman tests for multi-group comparisons, and Mann-Whitney U tests for pairwise analyses, with effect sizes reported as Cohen’s *d* and 95% confidence intervals used throughout to quantify uncertainty. The choice of non-parametric tests reflects the skewed, bounded nature of the outcome distributions and the use of the same 28 datasets across all conditions.

## 5. Results and Discussion

### 5.1. RQ1: Is LLM-guided Pseudo-Labelling a Reliable and Effective Approach for Abstract (Pre-)Screening?

RQ1 investigates whether LLM-guided pseudo-labelling can function as a reliable and effective approach to abstract pre-screening: whether the LLM ranking and the initial classifier it produces can identify relevant abstracts with sufficient reliability to constitute a useful, zero-annotation screening stage. Two complementary aspects of Stage S performance are examined: (a) the ranking quality of the LLM-generated document ordering, quantified through WSS@95%; and (b) the quality of the classifier trained exclusively on pseudo-labelled documents, assessed via Stage S success rate (≥95% recall at the 0.5 threshold of posterior probability), relaxed Stage S success rate (≥90% recall at the 0.5 threshold), recall (i.e., sensitivity), precision, F1-score, and AUC-ROC. The relaxed success rate is included to illustrate how close the End-of-Stage-S classifier comes to near-total relevant-document recovery across all datasets, capturing cases where recall falls just short of the strict 95% criterion. Because Stage S occurs before the active learning cycle, all results are independent of the query strategy in Stage A and reported collapsed across all six strategies.

#### 5.1.1. Stage S Ranking Quality and Classifier Performance across Ratios

Table 1 presents Stage S performance across all fourteen pseudo-labelling ratios and the random initialisation baseline (the setup in the original SAFE procedure). Under random initialisation, the End-of-Stage-A classifier achieves ≥95% recall on only 7.5% of datasets and ≥90% recall on only 10.0% of the tested datasets.

**Table 1.** End-of-Stage-S classifier quality and ranking performance: random initialisation versus all fourteen pseudo-labelling ratios (n = 28 datasets; 168 matched pairs per comparison). Δ = absolute improvement over random. *** *p* < 0.001 (Wilcoxon signed-rank). Coverage (%) = proportion of corpus pseudo-labelled (top *T*% + bottom *B*%). Stage S Success Rate (≥95%) = % of datasets with End-of-Stage-S classifier recall ≥95% at the 0.5 threshold; Relaxed Stage S Success (≥90%) applies the same criterion at 90%. Effect sizes for classifier metrics: Cohen’s *d* for Recall = 4.6 (large); *d* for AUC-ROC = 1.9-2.0 (large); *d* for F1 = 1.1-1.2 (large); *d* for Precision = 0.4 (small).

| Ratio | Stage S<br>Recall<br>(Mean±SD) | Stage S F1-Score<br>(Mean±SD) | Stage S<br>AUC-ROC<br>(Mean±SD) | Stage S<br>Precision<br>(Mean±SD) | WSS@95%<br>(Mean±SD) | Actual<br>WSS<br>(Mean±SD) | Stage S<br>Success<br>($\geq 95\%$) | Relaxed<br>Stage S<br>Success<br>($\geq 90\%$) |
| --- | --- | --- | --- | --- | --- | --- | --- | --- |
| Random | 0.136±0.187 | 0.051±0.083 | 0.640±0.151 | 0.064±0.083 | 0.199±0.240 | 0.041±0.061 | 7.5% | 10.0% |
| 2.5% | 0.994±0.004<br>$\Delta+0.858$*** | 0.133±0.020<br>$\Delta+0.082$*** | 0.903±0.016<br>$\Delta+0.263$*** | 0.078±0.013<br>$\Delta+0.014$*** | 0.620±0.246 | 0.294±0.140 | 96.4% | 96.4% |
| 5.0% | 0.998±0.001<br>$\Delta+0.862$*** | 0.143±0.022<br>$\Delta+0.092$*** | 0.909±0.013<br>$\Delta+0.269$*** | 0.084±0.015<br>$\Delta+0.020$*** | 0.646±0.225 | 0.342±0.131 | 100.0% | 100.0% |
| 7.5% | 0.998±0.001<br>$\Delta+0.862$*** | 0.147±0.022<br>$\Delta+0.096$*** | 0.905±0.013<br>$\Delta+0.265$*** | 0.087±0.015<br>$\Delta+0.023$*** | 0.633±0.217 | 0.365±0.128 | 100.0% | 100.0% |
| 10.0% | 0.997±0.002<br>$\Delta+0.861$*** | 0.152±0.023<br>$\Delta+0.101$*** | 0.910±0.012<br>$\Delta+0.270$*** | 0.090±0.015<br>$\Delta+0.026$*** | 0.637±0.214 | 0.386±0.126 | 100.0% | 100.0% |
| 12.5% | 0.996±0.002<br>$\Delta+0.861$*** | 0.157±0.024<br>$\Delta+0.106$*** | 0.909±0.011<br>$\Delta+0.269$*** | 0.094±0.016<br>$\Delta+0.030$*** | 0.642±0.208 | 0.404±0.117 | 96.4% | 100.0% |
| 15.0% | 0.996±0.002<br>$\Delta+0.861$*** | 0.159±0.024<br>$\Delta+0.108$*** | 0.908±0.011<br>$\Delta+0.268$*** | 0.096±0.017<br>$\Delta+0.032$*** | 0.642±0.208 | 0.412±0.106 | 96.4% | 100.0% |
| 17.5% | 0.995±0.003<br>$\Delta+0.860$*** | 0.160±0.025<br>$\Delta+0.109$*** | 0.906±0.011<br>$\Delta+0.266$*** | 0.097±0.017<br>$\Delta+0.033$*** | 0.636±0.199 | 0.418±0.099 | 96.4% | 100.0% |
| 20.0% | 0.994±0.003<br>$\Delta+0.859$*** | 0.160±0.025<br>$\Delta+0.109$*** | 0.904±0.010<br>$\Delta+0.264$*** | 0.097±0.017<br>$\Delta+0.033$*** | 0.626±0.196 | 0.419±0.094 | 96.4% | 100.0% |
| 22.5% | 0.993±0.003<br>$\Delta+0.858$*** | 0.162±0.025<br>$\Delta+0.111$*** | 0.901±0.010<br>$\Delta+0.261$*** | 0.099±0.018<br>$\Delta+0.035$*** | 0.625±0.188 | 0.425±0.088 | 96.4% | 96.4% |
| 25.0% | 0.992±0.004<br>$\Delta+0.857$*** | 0.163±0.025<br>$\Delta+0.112$*** | 0.899±0.009<br>$\Delta+0.259$*** | 0.100±0.018<br>$\Delta+0.036$*** | 0.626±0.178 | 0.429±0.084 | 92.9% | 96.4% |
| 27.5% | 0.992±0.026<br>$\Delta+0.856$*** | 0.165±0.171<br>$\Delta+0.114$*** | 0.895±0.066<br>$\Delta+0.255$*** | 0.101±0.122<br>$\Delta+0.037$*** | 0.621±0.177 | 0.435±0.080 | 92.9% | 96.4% |
| 30.0% | 0.991±0.026<br>$\Delta+0.855$*** | 0.168±0.174<br>$\Delta+0.117$*** | 0.892±0.067<br>$\Delta+0.252$*** | 0.103±0.124<br>$\Delta+0.039$*** | 0.622±0.166 | 0.444±0.077 | 92.9% | 96.4% |
| 32.5% | 0.990±0.029<br>$\Delta+0.854$*** | 0.168±0.176<br>$\Delta+0.117$*** | 0.890±0.068<br>$\Delta+0.250$*** | 0.104±0.127<br>$\Delta+0.040$*** | 0.622±0.162 | 0.446±0.070 | 92.9% | 96.4% |
| 35.0% | 0.990±0.029<br>$\Delta+0.854$*** | 0.170±0.178<br>$\Delta+0.119$*** | 0.887±0.068<br>$\Delta+0.247$*** | 0.105±0.129<br>$\Delta+0.041$*** | 0.619±0.161 | 0.451±0.066 | 92.9% | 96.4% |

Note. The relaxed success rate ( $\geq 90\%$ ) reaches 100% for ratios 5.0-20.0%, indicating that across these ratios every dataset achieves at least 90% End-of-Stage-S recall. The strict criterion ( $\geq 95\%$ ) is met by every dataset at three ratios (5.0%, 7.5%, 10.0%). The single dataset failing the strict criterion at 12.5-20.0% (CD012069) reaches 94.3-94.4% recall, one relevant document short of 95%. At 25.0-35.0%, two datasets fail the strict criterion: CD012069 (recall 0.869-0.883, falling below both strict and relaxed thresholds) and CD010558 (recall 0.919-0.946, failing only the strict criterion), accounting for the drop in strict success to 92.9% while relaxed success holds at 96.4%. Strict and relaxed success coincide at 2.5% (one dataset, CD012768, recalls only 86.0% at this ratio) and at 22.5% (both 96.4%). Random baseline averaged over 10 runs per dataset.

The contrast with random initialisation is stark across both metrics. All pseudo-labelling ratios achieve strict Stage S success rates of 92.9-100% compared to 7.5% under random initialisation (+85 to +93 pp), and relaxed Stage S success rates of 96.4-100% compared to 10.0% (+86to +90 pp). The relaxed success rate reveals that the single failing dataset at ratios 12.5-20.0% (CD012069) achieves recall just below the strict 95% threshold: it is recovering 94.4% of relevant studies from the End-of-Stage-S classifier alone, missing only one additional relevant document relative to the strict criterion. This illustrates that the strict failures are marginal and that the practical pre-screening performance across all 28 datasets is uniformly close to complete relevant-document recovery.

Strict Stage S success rate peaks at 5.0%, 7.5%, and 10.0% (100%), declines from 12.5% to 20.0% to 96.4%, and plateaus at 92.9% from 22.5% to 35.0%. The relaxed success rate remains 100% from 5.0% through 20.0%, drops to 96.4% at 22.5%, and remains stable at that level through 35.0%. While WSS@95% peaks at 5.0% (0.646) and remains broadly stable across ratios (0.619-0.646), actual WSS increases monotonically from 0.294 at 2.5% to 0.451 at 35.0%, reflecting improved classifier calibration with larger pseudo-labelled training sets.

Strict Stage S success rates remain at 92.9% (26/28) and relaxed success rates at 96.4% (27/28) across all four ratios. Two datasets account for all failures across this range. CD012069 falls below the relaxed 90% threshold for all ratios, and CD010558 falls below the strict 95% threshold but remains above 90%. Mean recall (0.990-0.992), F1-score (0.165-0.170), and AUC-ROC (0.887-0.895) remain stable, with AUC-ROC showing a gradual decline consistent with the trend observed from 22.5% onward, as pseudo-labelling progressively extends into less-confidently ranked boundary regions.

Table 1 establishes that pseudo-labelling reliably transforms the End-of-Stage-S classifier from near-useless to near-universally reliable abstract screener or pre-screener (92.9-100% strict, 96.4-100% relaxed) across all fourteen ratios. End-of-Stage-S classifiers at the ratios of 12.5-20.0% are, in practice, recovering most of the relevant studies on every benchmark dataset before any human annotation takes place.

#### 5.1.2. Discussion: RQ1?

The Stage S results raise four themes: a mechanistic explanation for near-perfect recall; interpreting the two success-rate metrics together; why the 5-10% range achieves 100% strict success; and the universality of strategy independence.

##### 5.1.2.1. Why Pseudo-Labelling Achieves Near-Perfect Recall

The ∼635% relative improvement in Stage S recall compared to random initialisation reflects a structural advantage created by the LLM ranking. Under random initialisation, the cold-start classifier encounters at most one or two positive training examples, producing unreliable probability estimates and low Stage S success rates. Pseudo-labelling reverses this: a strong ranking system tends to place more positive samples higher in the ranking, increasing the probability that pseudo-positives drawn from the top *T*% include a meaningful proportion of relevant documents and that pseudo-negatives drawn from the bottom *B*% are largely irrelevant. The classifier consequently learns from training data that is more representative of the positive class from the outset, addressing the cold-start problem at the level of training data composition. On the contrary, the more moderate AUC-ROC improvement (+42%) relative to recall (+635%) reflects the harder task of borderline discrimination, as the former measures ranking quality across all thresholds, including the difficult boundary region where the LLM’s scoring is imperfect.

##### 5.1.2.2. The Two Success Rate Metrics Read Together

The strict (≥95%) and relaxed (≥90%) Stage S success rates complement each other in revealing the distributional shape of Stage S failures. Where they coincide, as at 2.5% (both 96.4%) and 22.5% (both 96.4%), the failing dataset falls meaningfully short of the strict threshold, with recall dropping below 90%. At 2.5%, one dataset (CD012768) achieves only 86.0% Stage S recall, indicating that the pseudo-labelled training set is too sparse to reliably identify its relevant studies. At 22.5%, CD012069 similarly falls below 90%, suggesting that very aggressive pseudo-labelling may introduce excessive borderline noise, suppressing this dataset’s recall below the relaxed threshold.

Where strict and relaxed success rates diverge, as at 12.5-20.0% (strict 96.4%, relaxed 100%) and 25.0-35.0% (strict 92.9%, relaxed 96.4%), the datasets that fail the strict criterion still achieve above-90 % recall, indicating near misses rather than substantial failures. At 12.5-20.0%, the single failing dataset (CD012069) achieves 94.3-94.4% recalls, just short of the 95% threshold. At 25.0-35.0%, a second dataset (CD010558) additionally fails the strict criterion, achieving 91.9-94.6% recalls, while CD012069 drops below 90% and fails both thresholds. Based on the 28 benchmark datasets evaluated here, pseudo-labelling at 12.5-20.0% appears to support near-complete identification of relevant documents before any human annotation; however, whether this pattern generalises to reviews with different characteristics warrants further investigation.

##### 5.1.2.3. Why the 5-10% Range Achieves 100% Strict Success

The three ratios achieving 100% strict Stage S success (5.0%, 7.5%, and 10.0%) represent a zone in which the pseudo-labelled training set is large enough to provide reliable positive-class coverage yet small enough to avoid borderline noise that slightly degrades performance at higher ratios. At 5.0%, although only a small proportion of relevant documents may reside in the top 5% of the ranking, the pseudo-labelled training set provides the classifier with informative examples of what relevance looks like in the top-ranked documents and what irrelevance looks like in the bottom-ranked documents, enabling reliable classification from the outset. Above 10%, the bottom *T*% of pseudo-negatives have a higher chance of encountering borderline documents (harder samples). For datasets with particularly challenging prevalence characteristics (such as CD012069), this causes the End-of-Stage-S classifier to assign probabilities just below 0.5 to one or two relevant studies, resulting in a strict failure at the 95% threshold while remaining within the relaxed 90% threshold. The 5-10% range avoids both failure modes in our experiments: insufficient positive-class signal at very low ratios and borderline noise at very high ratios. Notably, this noise is a direct function of the ratio. As the pseudo-labelling proportion increases, the boundary between pseudo-positives and pseudo-negatives moves further from the extremes of the LLM ranking into a region of greater uncertainty, introducing progressively more mislabelled documents on both sides. Lower ratios concentrate pseudo-labels at the most confidently ranked positions, where labelling error is lowest; higher ratios extend into the middle of the ranking, where the LLM’s discrimination is weakest, and label noise is highest.

The divergence between the Stage S success rates (peaking at 5-10%) and the Stage A success rates reported in RQ3 (which increase monotonically with the ratio up to 22.5%) reflects different mechanisms. Stage S success depends on the 0.5-threshold decision boundary being correctly calibrated, which moderate pseudo-labelling achieves most reliably: at the 5.0-10.0% ratios, the default 0.5 threshold already yields a 96% Stage S success rate. Therefore, threshold calibration in the manner of Wang et al. (2024), who tuned the threshold post hoc against a predefined recall target for zero-shot LLM screening, offers a further reliability lever. Stage A success depends on the active learning classifier eventually finding all relevant documents, which benefits from a larger initial training set, even at the cost of some borderline noise in the pseudo-labelled negatives. These results suggest that the 5-10% range is most effective for Stage S pre-screening reliability. At the same time, higher ratios may provide greater downstream benefits in the full active learning pipeline. However, this trade-off should be interpreted in the context of the 28 datasets examined here. Very large-scale benchmarking is planned as our future work.

### 5.2. RQ2: What Is the Safest Configuration for Pseudo-Labelling towards Reliable Abstract Screening?

RQ2 identifies the pseudo-labelling configuration that maximises screening reliability, operationalised as the Stage A success rate, i.e. the success rate of the End-of-Stage-A classifier. Two design choices are evaluated: the pseudo-labelling ratio and the active learning query strategy. Their independent effects on success rate and WSS are tested using Friedman tests, with the joint configuration that maximises Stage A reliability identified subsequently.

#### 5.2.1. Safest Pseudo-Labelling Ratio

Table 2 ranks all 14 pseudo-labelling ratios by strict success rate. The top positions are held by ratios between 22.5% and 35.0%, all achieving strict success rates of 94.6% or higher. Ratios of 22.5% and 30.0% are jointly ranked first, each achieving a strict success rate of 95.8% across 168 experiments (28 datasets × 6 strategies). The relaxed success rate (≥90%) reveals that many experiments falling just short of the strict criterion are near-misses, particularly at lower ratios where the strict-relaxed gap is widest.

**Table 2.** Pseudo-labelling ratios ranked by strict success rate (*n* = 28 datasets, all six query strategies; 168 experiments per ratio). Strict Success = % achieving ≥95% total recall. Relaxed Success = % achieving ≥90% total recall. Gap = relaxed − strict (pp).

| Rank | Ratio | Strict Success ($\geq 95\%$) | Relaxed Success ($\geq 90\%$) | Gap (pp) | Mean WSS (Stage A) | Mean Recall (%) |
| --- | --- | --- | --- | --- | --- | --- |
| 1 | **22.5%** | **95.8%** | 98.2% | 2.4 | 0.429 | 99.1% |
| 2 | **30.0%** | **95.8%** | 98.8% | 3.0 | 0.390 | 99.3% |
| 3 | **27.5%** | **95.2%** | 99.4% | 4.2 | 0.407 | 99.3% |
| 4 | 25.0% | 94.6% | 98.2% | 3.6 | 0.414 | 99.1% |
| 5 | 32.5% | 94.6% | 97.6% | 3.0 | 0.396 | 99.2% |
| 6 | 35.0% | 94.6% | 98.2% | 3.6 | 0.391 | 99.3% |
| 7 | 20.0% | 91.7% | 98.8% | 7.1 | 0.434 | 98.9% |
| 8 | 15.0% | 90.5% | 97.0% | 6.5 | 0.459 | 98.5% |
| 9 | 17.5% | 89.3% | 96.4% | 7.1 | 0.462 | 98.6% |
| 10 | 12.5% | 86.3% | 92.3% | 6.0 | 0.490 | 97.6% |
| 11 | 10.0% | 82.1% | 89.3% | 7.2 | 0.514 | 96.9% |
| 12 | 7.5% | 78.0% | 88.1% | 10.1 | 0.522 | 95.6% |
| 13 | 5.0% | 72.0% | 84.5% | 12.5 | 0.550 | 94.0% |
| 14 | 2.5% | 58.3% | 75.0% | 16.7 | 0.509 | 90.5% |

Note. The Gap column shows the difference in percentage points between the relaxed and strict success rates, representing experiments in which total recall falls between 90% and 95%. The gap is largest at lower ratios (2.5%: +16.7pp) and narrows considerably at higher ratios (22.5%: +2.4pp; 30.0%: +3.0pp), confirming that higher ratios not only achieve more successes but also produce fewer near-miss cases.

The relaxed success rate reveals an important characteristic of lower pseudo-labelling ratios: at 2.5%, while strict success is only 58.3%, a further 16.7 percentage points (pp hereafter) of experiments (75.0% relaxed) achieve at least 90% recall, meaning many experiments at low ratios are near-misses rather than outright failures. As the ratio increases, the gap narrows substantially. At 20.0%, only 7.1pp separates relaxed (98.8%) and strict (91.7%) success; at 22.5%, the gap shrinks to 2.4pp (95.8% strict, 98.2% relaxed); and at 30.0%, the gap is 3.0pp (95.8% strict, 98.8% relaxed). Notably, 27.5% achieves the highest relaxed success (99.4%) of all ratios while also placing in the top three for strict success (95.2%). This convergence at higher ratios confirms that pseudo-labelling beyond approximately 20% not only achieves stricter success rates but also converts near-misses into complete successes, yielding more reliable and consistent outcomes overall.

#### 5.2.2. Size-Adaptive Safest Ratios

Table 3 presents size-adaptive recommendations based on the highest achievable strict success rate at any ratio for each dataset size category, and Figure 1 shows the full strict and relaxed success-rate curves across all ratios, stratified by size.

**Figure 1.**
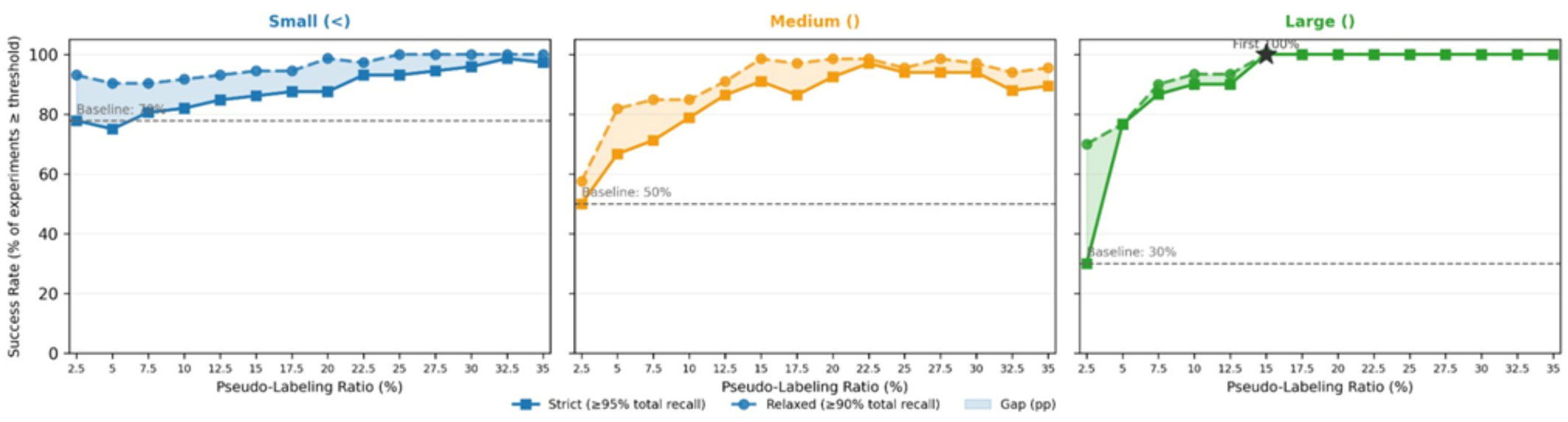
Stage A success rate as a function of pseudo-labelling ratio, stratified by dataset size. Solid lines with squares = strict criterion (≥95% total recall); dashed lines with circles = relaxed criterion (≥90% total recall). Shaded regions = near-miss gap between relaxed and strict thresholds. Dashed horizontal lines = random initialisation, strict baseline per size category. Stars mark the first ratio where the strict success rate reaches 100%.

**Table 3.** Size-adaptive safest pseudo-labelling ratios by dataset size category. Success rates are averaged across all six query strategies at the stated optimal ratio.

| Size Category | N | Safest Ratio | Strict Success Rate ($\geq 95\%$) | Relaxed Success Rate ($\geq 90\%$) | Mean WSS (Stage A) |
| --- | --- | --- | --- | --- | --- |
| Small (<1,000) | 12 | 32.5% | 98.6% | 100.0% | 0.294 |
| Medium (1K–5K) | 11 | 22.5% | 97.0% | 98.5% | 0.492 |
| Large (>5,000) | 5 | 15.0% | 100.0% | 100.0% | 0.595 |

Note. The safest ratio varies markedly across size categories, reflecting the inverse relationship between dataset size and the ratio required for reliable initialisation. For large datasets, strong recall signals allow reliable active learning at a lower ratio (15.0%), achieving perfect strict and relaxed success. Small datasets require a substantially higher ratio (32.5%) to approach 98.6% strict success, as the low absolute number of relevant documents creates greater classification uncertainty at any given initialisation size.

The size-stratified curves reveal a coherent set of trajectories rather than three qualitatively different ones. Larger datasets reach high success at lower pseudo-labelling ratios, whereas smaller datasets require higher ratios to seed the active learning cycle with a sufficient absolute number of positives. **Large datasets (green, n = 5)** rise most steeply from a baseline of 30%: relaxed success is near-perfect by 7.5%, and strict success reaches 100% at 15.0% and remains there at all higher ratios. **Medium datasets (orange, n = 11)** follow the same general shape from a baseline of 50%, requiring a slightly larger ratio before plateauing, with strict success settling between 90% and 97% above 15.0%. In both of these categories, the strict-relaxed gap is widest at the lower ratios and narrows progressively as the ratio increases, indicating that near misses become rarer with stronger initialisation. **Small datasets (blue, n = 12)** depart from this pattern. They begin from the highest random baseline (78%), reflecting the relative simplicity of smaller corpora, yet improve only gradually, with strict success approaching 100% only above 30.0%. More informatively, the strict and relaxed curves run consistently close to each other across the entire range, indicating that almost all strict failures in the small-dataset regime are marginal near misses (recall in [90%, 95%)) rather than outright failures. The clear cross-size ordering, large leading, medium leading, and small, holds at all ratios and motivates the size-adaptive recommendations in Table 3.

#### 5.2.3. Safest Query Strategy

Table 4 ranks the six query strategies by strict success rate, averaged across all 14 pseudo-labelling ratios (2.5%–35%), providing an overall reliability ranking independent of any specific ratio choice. Pure Uncertainty leads by a substantial margin, achieving a mean strict success rate of 95.4% and relaxed success rate of 99.2%, confirming its dominance regardless of the ratio. All strategies achieve relaxed success rates above 80% on average, but strategies differ considerably in their strict success rates (81.1%–95.4%) and near-miss gaps (3.8-9.2pp).

**Table 4.**
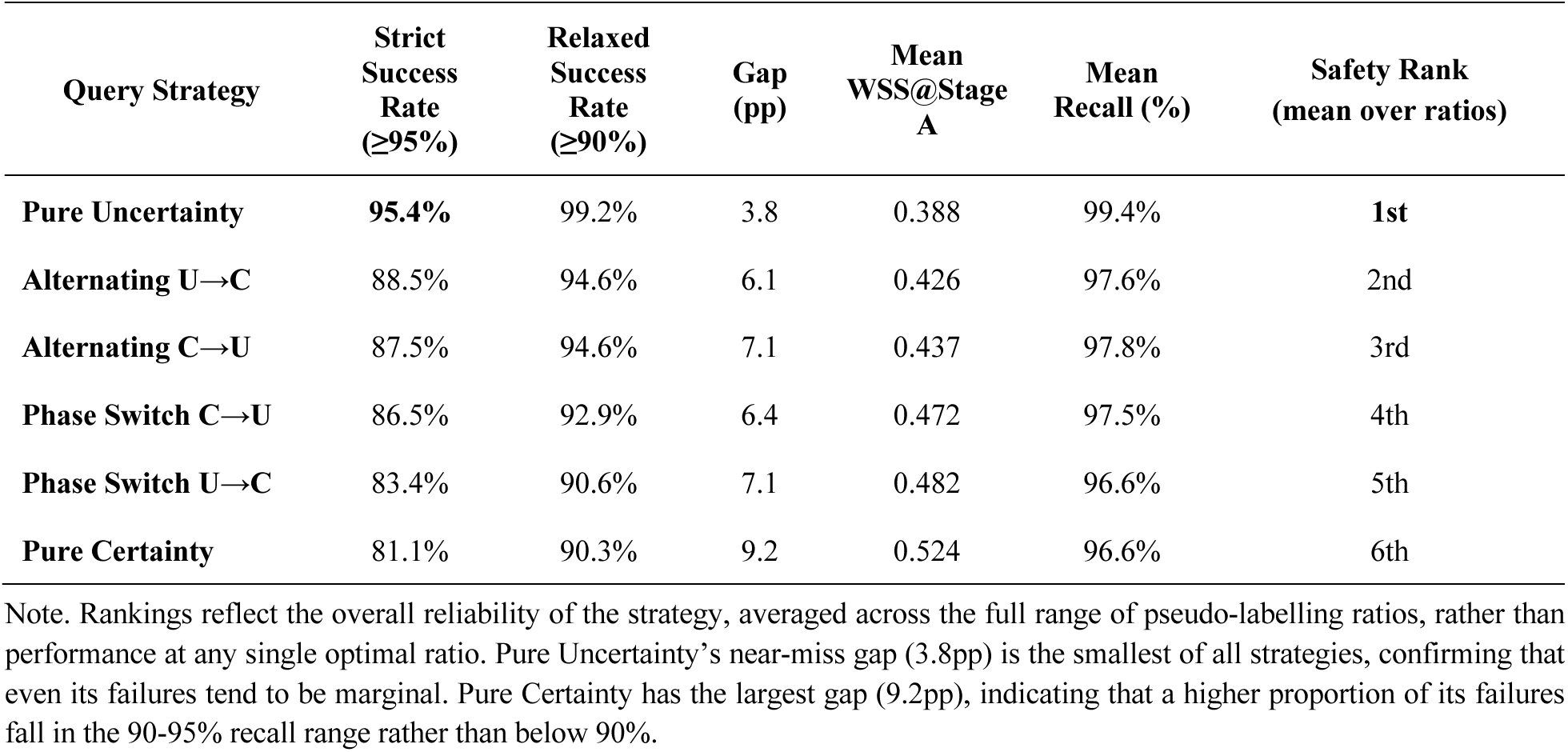
Query strategy overall performance ranked by strict success rate, averaged across all 14 pseudo-labelling ratios (2.5%–35%; 14 × 28 = 392 experiments per strategy). Strict SR = % achieving ≥95% total recall. Relaxed SR = % achieving ≥90% total recall. Gap = relaxed − strict (pp). Mean WSS and recall are averaged across all ratios and datasets.

Pure Uncertainty’s mean strict success of 95.4% is 6.9pp higher than the next-best strategy (Alternating U→C at 88.5%), and its near-miss gap of 3.8pp is the smallest of all six strategies. This confirms that Pure Uncertainty is not only the most reliable strategy on average but also the most consistent: when it fails to meet the strict threshold, the shortfall is typically small. However its reliability is at the cost of lower WSS. The four hybrid strategies cluster between 83.4% and 88.5% strict success with gaps of 6.1-7.1pp, offering a meaningful trade-off of somewhat lower reliability in exchange for higher work savings (WSS 0.426-0.482 vs 0.388 for Pure Uncertainty). Pure Certainty ranks last (81.1% strict, gap 9.2pp), confirming that its exploitation-focused mechanism is structurally less suited to the strict ≥95% recall criterion across varying ratio conditions.

#### 5.2.4. Safest Configurations: Full Strategy (Ratio Heatmap)

Figure 2 presents the full heatmap of strict success rates across all 84 strategy-ratio combinations (6 strategies × 14 ratios), with the right panel showing the mean WSS@Stage A for the subset of configurations that achieve 100% strict success across all 28 datasets (starred cells). Ten configurations achieve 100% strict success across all 28 datasets, marked with asterisks in Figure 2. The right panel of Figure 2 shows the mean WSS@Stage A for four confirmed 100%-success configurations: Phase Switch U→C at 30.0% achieves the highest work savings (WSS = 0.446), followed by Pure Uncertainty at 12.5% and Alternating U→C at 22.5% (both WSS = 0.407), and Alternating C→U at 22.5% (WSS = 0.406). The heatmap reveals a broad region of high success rates spanning moderate-to-high pseudo-labelling ratios across most query strategies, rather than a single privileged configuration. Two row-level patterns are robust to dataset-level noise: Pure Uncertainty reaches the ceiling at lower ratios than the other strategies, and Pure Certainty trails throughout. Beyond these, the heatmap does not support stronger conclusions about individual (ratio, strategy) cells. With n = 28 datasets, a 100% success rate (28/28) and a 96.4% rate (27/28) differ by exactly one dataset, so the asterisked configurations are best read as falling within rounding distance of the maximum rather than as categorically superior to their neighbours. The same caution applies to the work-saved comparison: the four top configurations cluster within 0.04 WSS of each other (0.406-0.446), a margin too narrow to support a firm ranking given dataset-level variability. The defensible conclusion is therefore that high reliability and meaningful work savings are jointly attainable across a broad stable region of the configuration space, not that any specific (ratio, strategy) combination is uniquely optimal.

**Figure 2.**
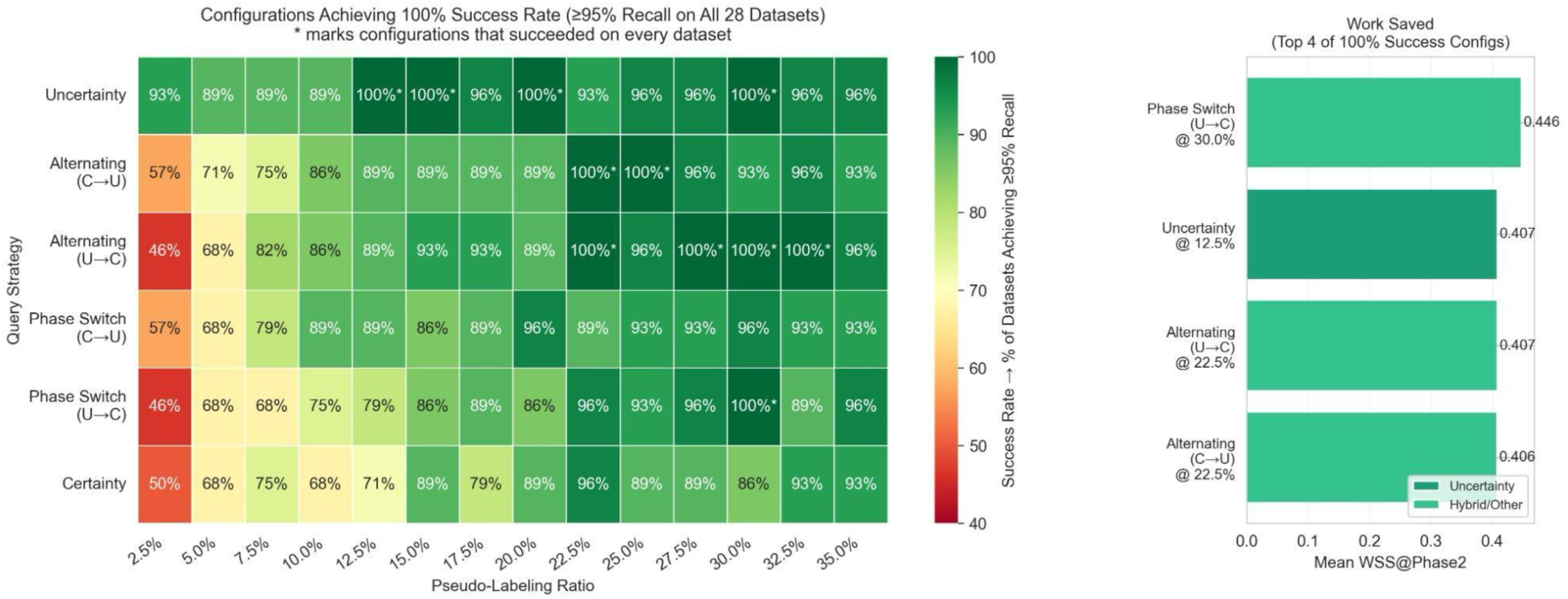
Strict Stage A success rate (% of 28 datasets achieving ≥95% total recall) for all 84 strategy-ratio configurations. Asterisks (*) mark configurations that succeeded on every dataset. Right panel: mean WSS@Stage A for the top 4 confirmed 100%-success configurations (Phase Switch U→C at 30.0%, Pure Uncertainty at 12.5%, Alternating U→C at 22.5%, Alternating C→U at 22.5%), showing the efficiency trade-off across these optimal points. Colour scale: dark green = 100%, red = ≤50%.

#### 5.2.5. Strategy Performance by Dataset Size

Table 5 reveals that Pure Uncertainty is uniquely robust across all dataset sizes when averaged over all ratios: it is the only strategy achieving 100% strict and relaxed success on large datasets (mean over all ratios), and it leads the ranking on both small and medium datasets (96.4% and 92.2% strict, respectively). On large datasets, the performance hierarchy is clearer: Pure Uncertainty achieves perfect success (100% strict), while all other strategies fall to 85.7-91.4% strict success on average, reflecting that even strong strategies occasionally fail on at least one large dataset when low pseudo-labelling ratios are included in the average. On medium datasets, the Alternating strategies (87.7% and 84.4% strict) closely follow Pure Uncertainty (92.2%), while Phase Switch U→C and Pure Certainty drop to 78-79% strict, driven by poor performance at low ratios. The consistent WSS gradient and lower WSS for strategies that rank higher on reliability confirm a fundamental trade-off: strategies that prioritise exploration (Pure Uncertainty) achieve higher recall at the cost of reviewing more documents, while exploitation-heavy strategies (Pure Certainty) save more work but miss more reviews.

**Table 5.** Strategy strict and relaxed success rate (%) and mean WSS@Stage A by dataset size category, averaged across all 14 pseudo-labelling ratios. Small = <1,000 records (*n* = 12); Medium = 1000-5000 (*n* = 11); Large = >5000 (*n* = 5).

| Strategy | Small<br>Strict% | Small<br>Relaxed% | Small<br>WSS | Med.<br>Strict% | Med.<br>Relaxed% | Med.<br>WSS | Large<br>Strict% | Large<br>Relaxed% | Large<br>WSS |
| --- | --- | --- | --- | --- | --- | --- | --- | --- | --- |
| **Pure Uncertainty** | 96.4% | 99.4% | 0.243 | 92.2% | 98.7% | 0.464 | 100.0% | 100.0% | 0.568 |
| **Alternating U→C** | 88.1% | 95.8% | 0.310 | 87.7% | 93.5% | 0.484 | 91.4% | 94.3% | 0.577 |
| **Alternating C→U** | 88.7% | 95.2% | 0.308 | 84.4% | 92.9% | 0.511 | 91.4% | 97.1% | 0.588 |
| **Phase Switch C→U** | 86.3% | 94.6% | 0.345 | 84.4% | 90.3% | 0.550 | 91.4% | 94.3% | 0.606 |
| **Phase Switch U→C** | 86.9% | 95.2% | 0.343 | 78.6% | 85.7% | 0.563 | 85.7% | 90.0% | 0.637 |
| **Pure Certainty** | 82.1% | 95.2% | 0.418 | 77.9% | 84.4% | 0.596 | 85.7% | 91.4% | 0.618 |

Note. All figures represent averages across all 14 ratios (2.5%–35%), reflecting overall strategy reliability rather than performance at any single ratio. Pure Uncertainty is the only strategy achieving 100% strict and relaxed success on large datasets across the full ratio range. On small and medium datasets, no strategy achieves 100% strict success on average, as lower ratios reduce success rates; this is expected and motivates the size-adaptive ratio recommendations in Table 3.

#### 5.2.6. Recommended Safest Configurations

In answer to RQ2, the safest universal ratios, jointly achieving 95.8% strict success across all strategies and datasets, are 22.5% and 30.0%. Overall, 22.5%-35% seems to be the safer region, particularly for the two Alternating strategies while Uncertainty is the strategy that is less sensitive to pseudo-labelling ratio when the pseudo-labelling ratio is sufficiently large, say ≥12.5% (Figure 2). When the dataset size is known, size-adaptive ratios of 32.5% (small), 22.5% (medium), and 15.0% (large) yield strict success rates of 98.6%, 97.0%, and 100.0%, respectively. At the strategy level, Pure Uncertainty is the safest overall, achieving a mean strict success rate of 95.4% across all ratios, more than 6pp ahead of the next-best strategy. Ten strategy-ratio configurations achieved 100% strict success across all 28 datasets, with the top four (Phase Switch U→C at 30.0%, Pure Uncertainty at 12.5%, Alternating U→C at 22.5%, and Alternating C→U at 22.5%) shown in Figure 2. Among these, Pure Uncertainty at 12.5% is the most ratio-efficient, achieving complete reliability while pseudo-labelling the smallest fraction of the corpus. The cost saving here is computational rather than financial: a smaller pseudo-labelled set reduces the memory footprint of the labelled training pool held by the active learning classifier and shortens the time required to train and re-train it across active learning iterations (WSS = 0.407). Across all size categories, Pure Uncertainty is the only strategy achieving 100% strict success on large datasets when averaged over all ratios, confirming its unique universality.

#### 5.2.7. Discussion: RQ2

The RQ2 results raise four interpretive themes: why higher ratios are safer; why Pure Uncertainty achieves 100% success at the lowest ratio; why certainty-based strategies have structural limitations even at their optimal ratio; and the practical significance of the 100%-success configurations and size-adaptive recommendations.

##### 5.2.7.1. Why Higher Ratios Are Safer

The broadly monotonic relationship between ratio and strict success rate (58.3% at 2.5% to 95.8% at 22.5-30.0%) is not simply a function of having more training examples. As established in RQ1, Stage S recall is near-perfect across all ratios, confirming that the LLM ranking already recovers nearly all relevant documents regardless of the number of pseudo-labels drawn. The safety gains from higher ratios therefore operate through a different mechanism: the composition of the pseudo-labelled training set changes as the ratio increases.

Because pseudo-labels are drawn from an LLM-ranked list that is imperfect, higher ratios incorporate proportionally more mislabelled examples. Average pseudo-label noise increases monotonically with ratio, from 0.281 at 2.5% to 0.429 at 35.0%. Rather than degrading performance, this increasing noise appears to be beneficial: it prevents the End-of-Stage-A classifier from overfitting to the most confidently ranked, typically easier positive examples near the top of the LLM ranking, and instead forces a more lenient decision boundary that remains sensitive to difficult positives located deeper in the ranking. At low ratios, the pseudo-labelled set is dominated by high-confidence LLM predictions, producing a sharp but potentially over-specialised boundary that may exclude atypical or weakly-signalled relevant documents. At higher ratios, the boundary broadens and becomes more tolerant, reducing the likelihood of premature convergence before all relevant studies are found.

The relaxed success rate adds nuance to this picture. At 2.5%, the gap between strict (58.3%) and relaxed (75.0%) success is 16.7pp, meaning roughly one in six experiments achieves 90-95% recall, a near-miss rather than an outright failure. At 22.5%, the gap narrows to 2.4pp. At 27.5%, the relaxed rate (99.4%) exceeds that of any other ratio, suggesting this range represents a near-optimal point where the classifier boundary is broad enough to capture difficult positives without being so noisy as to degrade ranking quality. Beyond 30.0%, marginal gains in strict success diminish while WSS declines slightly, suggesting that the additional noise above approximately 25-30% begins to dilute the classifier signal without further improving boundary coverage, yielding a diminishing-returns regime.

##### 5.2.7.2. Why Pure Uncertainty Achieves 100% Success at the Lowest Ratio

Pure Uncertainty achieves a mean strict success rate of 95.4% across all 14 ratios, 6.9pp ahead of the next-best strategy (Alternating U→C), and is the only strategy achieving 100% strict and relaxed success on large datasets when averaged over the full ratio range. Its boundary-exploration mechanism, which systematically samples documents with the highest classifier uncertainty, ensures near-complete coverage of all relevant sub-regions of documents before the SAFE stopping criterion is triggered. This makes it particularly robust to imperfect initial ranking: when pseudo-labels do not perfectly separate relevant from irrelevant documents, active learning under Pure Uncertainty will seek out under-sampled regions, including clusters of relevant documents that a weaker initial classifier may have ranked near the boundary. Critically, this robustness persists across all ratio conditions: when its strict success rate is averaged over all 14 pseudo-labelling ratios, Pure Uncertainty exceeds every other strategy. In other words, it maintains its advantage at low ratios (where initialisation quality is lower) and at high ratios (where all strategies perform well), resulting in the consistently highest average strict success rate across the benchmark, at the sacrifice of WSS (Table 5) and convergence speed (RQ4).

##### 5.2.7.3. Why Certainty-Based Strategies Have Structural Limitations Even at Their Optimal Ratio

Pure Certainty underperforms across the entire ratio range, never reaching 100% strict success at any single ratio in Figure 2 and plateauing at 93% (26/28 datasets) at the highest ratios. Its mean strict success rate of 81.1% is the lowest of the six strategies, and its near-miss gap of 9.2pp is the largest, indicating that a meaningful proportion of failures fall outside even the relaxed 90% threshold. These limitations persist regardless of ratio and are therefore structural rather than configurational.

The mechanism is straightforward. Certainty-based sampling preferentially queries documents the classifier is most confident are relevant, which under pseudo-labelled initialisation cluster tightly around the prototypical positives identified by the LLM ranking. This produces rapid early discovery of relevant documents but leaves the long tail of atypical or weakly-signalled positives under-sampled. As the high-confidence pool is exhausted, the classifier’s top-ranked predictions increasingly include borderline cases, and the SAFE criterion of no relevant records in the last 50 consecutive screenings can fire while a residual fraction of harder positives remains undiscovered. The 9.2pp near-miss gap reflects precisely this: missing recall typically sits in the 90-95% band rather than below 90%.

This pattern bears on prior conclusions about certainty-based screening. Miwa et al. (2014) argued, on the basis of simulation experiments, that certainty-based sampling reduces screening workload relative to uncertainty-based alternatives. Their analysis, like much of the subsequent simulation literature, evaluated strategies through theoretical convergence metrics such as the proportion of the corpus screened to reach a fixed recall threshold. The present results do not contradict that finding under those metrics: in RQ4, Pure Certainty under pseudo-labelled initialisation does converge faster on % Screened to 95% Recall. The difficulty is that such metrics are oracle measures, requiring advance knowledge of the total number of relevant documents, which is the very quantity that abstract screening seeks to determine. A strategy that performs well under oracle-based metrics is not guaranteed to perform well under any operational stopping rule that must decide when to stop without that knowledge.

The RQ3 results make this disconnect concrete. Under SAFE Stage A stopping, Pure Certainty achieves only 81.1% mean strict success across ratios and never reaches the 100% ceiling attained by Pure Uncertainty, the Alternating strategies, or Phase Switch U→C. The same mechanism that produces fast theoretical convergence is what makes the strategy vulnerable to premature stopping. Conclusions about strategy preference drawn from theoretical convergence simulations should therefore be interpreted with caution when the strategy is to be deployed under an operational stopping criterion, a concern that motivates the argument developed more fully in RQ4.

##### 5.2.7.4. Practical Significance of the 100%-Success Configurations and Size-Adaptive Recommendations

The ten 100%-success configurations demonstrate that the cold-start problem can be fully resolved across all 28 benchmark datasets at multiple strategy-ratio combinations. The right panel of Figure Y highlights the top four such configurations: Phase Switch U→C at 30.0% achieves the highest WSS (0.446), followed by Pure Uncertainty at 12.5% (WSS 0.407), and both Alternating strategies at 22.5% (WSS 0.406-0.407). The WSS ordering among these four configurations quantifies the efficiency-ratio trade-off: practitioners can achieve the same complete reliability (100% strict success) with a lower ratio under Pure Uncertainty (12.5%) than under any hybrid strategy, but if a hybrid strategy is preferred, Alternating variants at 22.5% offer an equivalent reliability guarantee with marginally lower work savings.

The size-adaptive recommendations (32.5% for small, 22.5% for medium, 15.0% for large) operationalise a key empirical pattern in the data: larger datasets with richer evidence signals require less pseudo-labelling to achieve reliable initialisation. For large datasets, 15.0% is sufficient to reach 100% strict and relaxed success; for medium datasets, 22.5% approaches 97-99% strict success; and small datasets require ratios above 30.0% for near-perfect strict success. Practitioners who know their corpus size before screening can use these thresholds to avoid over-labelling large datasets or under-labelling small ones. These recommendations are established on the CLEF eHealth 2019 benchmark and should be validated before assuming universal applicability to reviews with substantially different prevalence rates or corpus structures.

### 5.3. RQ3: Can LLM-based Pseudo-Labelling Enhance the SAFE Procedure to Achieve Reliable and More Effective Abstract Screening?

RQ3 evaluates whether pseudo-labelled initialisation translates the Stage S gains established in RQ1 into improved end-to-end screening outcomes in the full SAFE pipeline (in our case stopping at the end of Stage A). Whereas RQ1 assessed the End-of-Stage-S classifier in isolation and RQ2 identified the safest configuration for Stage A, RQ3 examines whether these upstream improvements propagate through Stage A active learning to produce higher total recall, greater work savings, and a lower proportion of records screened by the time the SAFE stopping criteria are met. Three initialisation conditions are evaluated: Universal 22.5% (the universal ratio identified as jointly safest in RQ2), Fixed 30.0% (a second high-performing fixed ratio), and Optimal-by-Size (size-adaptive ratios: 32.5% for small datasets, 22.5% for medium, and 15.0% for large), each compared to the random initialisation baseline of the original SAFE procedure. Comparisons are conducted across all 28 CLEF eHealth 2019 datasets and all six query strategies (168 matched triplets per condition), with statistical testing via Wilcoxon signed-rank tests and effect sizes reported as Cohen’s *d*.

#### 5.3.1. Overall Final Screening Outcomes

Table 6 summarises end-to-end screening performance for each initialisation condition, averaged across all strategies and datasets. Figure 3 presents the same four metrics as grouped bar charts, illustrating the magnitude of the improvement over random initialisation.

**Figure 3.**
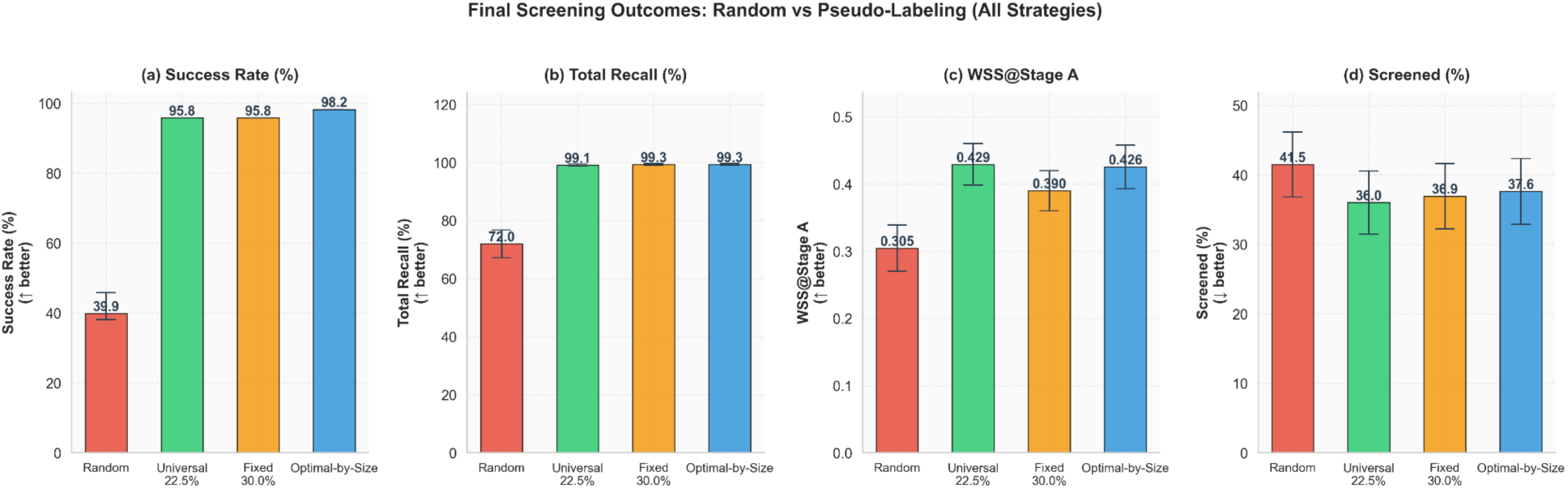
Final screening outcomes under random initialisation and three pseudo-labelling conditions (Universal 22.5%, Fixed 30.0%, Optimal-by-Size), averaged across all six query strategies and 28 datasets. (a) Success Rate (≥95% total recall); (b) Total Recall (%); (c) WSS@Stage A; (d) Screened (%). Error bars = ±1 SD (not shown for Success Rate, which is a proportion).

**Table 6.**
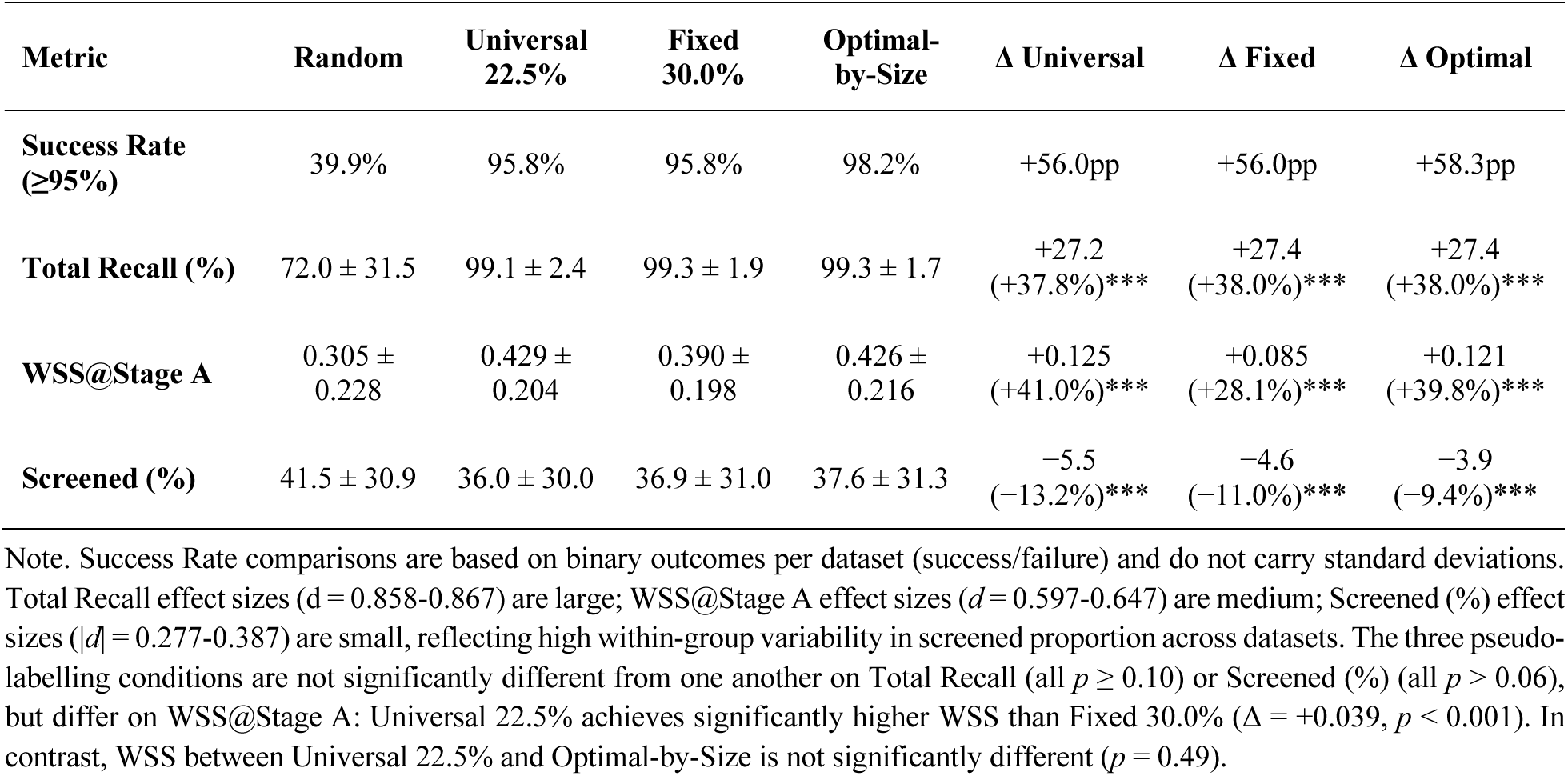
Final screening outcomes under random initialisation versus three pseudo-labelling conditions, averaged across all six query strategies (*n* = 28 datasets; 168 observations per condition). Success Rate = % of datasets achieving ≥95% total recall. Total Recall and Screened (%) are reported as Mean ± SD. *** *p* < 0.001 (Wilcoxon signed-rank). Δ columns show absolute improvement relative to the random baseline. Cohen’s *d*: |*d*| ≥ 0.8 = large, 0.5-0.8 = medium, 0.2-0.5 = small.

**Table 7.** Final screening outcomes by query strategy and initialisation condition (n = 28 datasets per strategy). Success Rate = % of datasets achieving ≥95% total recall. WSS@Stage A = mean work saved over sampling at Stage A completion.5% or Optimal-by-Size achieve 100% success rate for that strategy.

| Query Strategy | Success Rate (%) |  |  |  | WSS@Stage A |  |  |  |
| --- | --- | --- | --- | --- | --- | --- | --- | --- |
|  | Random | Univ.<br>22.5% | Fixed<br>30.0% | Opt.-by-Size | Random | Univ.<br>22.5% | Fixed<br>30.0% | Opt.-by-Size |
| **Pure Uncertainty** | 35.7% | 92.9% | 100.0% | 96.4% | 0.254 | 0.371 | 0.367 | 0.368 |
| **Alternating U→C** | 39.3% | 100.0% | 100.0% | 100.0% | 0.325 | 0.407 | 0.352 | 0.406 |
| **Alternating C→U** | 39.3% | 100.0% | 92.9% | 100.0% | 0.322 | 0.406 | 0.331 | 0.399 |
| **Phase Switch U→C** | 50.0% | 96.4% | 100.0% | 96.4% | 0.278 | 0.474 | 0.446 | 0.463 |
| **Phase Switch C→U** | 39.3% | 89.3% | 96.4% | 96.4% | 0.321 | 0.445 | 0.391 | 0.446 |
| **Pure Certainty** | 35.7% | 96.4% | 85.7% | 100.0% | 0.327 | 0.471 | 0.454 | 0.471 |

Note. All figures are computed across 28 datasets for each strategy-initialisation combination. The Random condition reflects ten averaged independent runs per dataset. Fixed 30.0% achieves 100% success for Pure Uncertainty, Alternating U→C, and Phase Switch U→C, but underperforms Universal 22.5% for Alternating C→U (92.9% vs 100%) and Pure Certainty (85.7% vs 96.4%). Optimal-by-Size achieves 100% success for Pure Certainty and both Alternating strategies while matching or exceeding other conditions on all remaining strategies.

The improvement associated with pseudo-labelled initialisation is large and consistent across all four outcome metrics. Under random initialisation, only 39.9% of the 168 strategy-dataset combinations achieve ≥95% total recall; under Universal 22.5% and Fixed 30.0%, this proportion rises to 95.8% in both cases (+56.0pp); and under Optimal-by-Size, to 98.2% (+58.3pp). These improvements represent a near-complete resolution of the reliability deficit that characterises random initialisation in the original SAFE procedure.

The total recall gains are statistically robust and practically meaningful. Recall increases from a mean of 72.0% under random initialisation to 99.1-99.3% across the three pseudo-labelling conditions (all *p* < 0.001, Cohen’s *d* = 0.858-0.867, significant with large effects). The negligible differences among the three pseudo-labelling conditions in total recall (all pairwise *p* ≥ 0.10) confirm that, once pseudo-labelled initialisation is applied, total recall converges to near-ceiling levels regardless of whether a fixed or size-adaptive ratio is selected.

Work savings also improve substantially. WSS@Stage A increases from 0.305 (random) to 0.429 under Universal 22.5% (a relative 41.0% increase, Cohen’s *d* = 0.647, medium effect), 0.390 under Fixed 30.0% (a relative 28.1% increase, Cohen’s *d* = 0.415, small effect), and 0.426 under Optimal-by-Size (a relative 39.8% increase, Cohen’s *d* = 0.597, medium effect). Here, the three conditions are not equivalent: Universal 22.5% achieves statistically significantly higher WSS than Fixed 30.0% (Δ = +0.039, *p* < 0.001), despite both achieving the same success rate. Optimal-by-Size occupies an intermediate position, with WSS (0.426) statistically indistinguishable from Universal 22.5% (*p =* 0.49) but significantly higher than Fixed 30.0% with statistical significance (Δ = +0.036, *p* < 0.001).

The proportion of records screened by study completion decreases from 41.5% (random) to 36.0% (Universal 22.5%), 36.9% (Fixed 30.0%), and 37.6% (Optimal-by-Size), all statistically significant reductions (*p* < 0.001, *p <* 0.001*, p* < 0.001, respectively). Effect sizes are small (|*d*| = 0.277-0.387), reflecting high variance in the proportion screened across datasets. Still, the consistent direction across all three conditions confirms that pseudo-labelled initialisation reliably reduces the screening burden even when averaged over a diverse set of review sizes and topic types.

#### 5.3.2. Dataset-Level Improvements: Scatter Analysis

Figure 4 presents pairwise scatter plots comparing total recall, WSS@Stage A, and proportion screened (i.e., % Screened) between the random baseline and the two fixed pseudo-labelling conditions (Universal 22.5% and Fixed 30.0%) across all 168 strategy-dataset pairs. Each point corresponds to one strategy-dataset combination.

**Figure 4.**
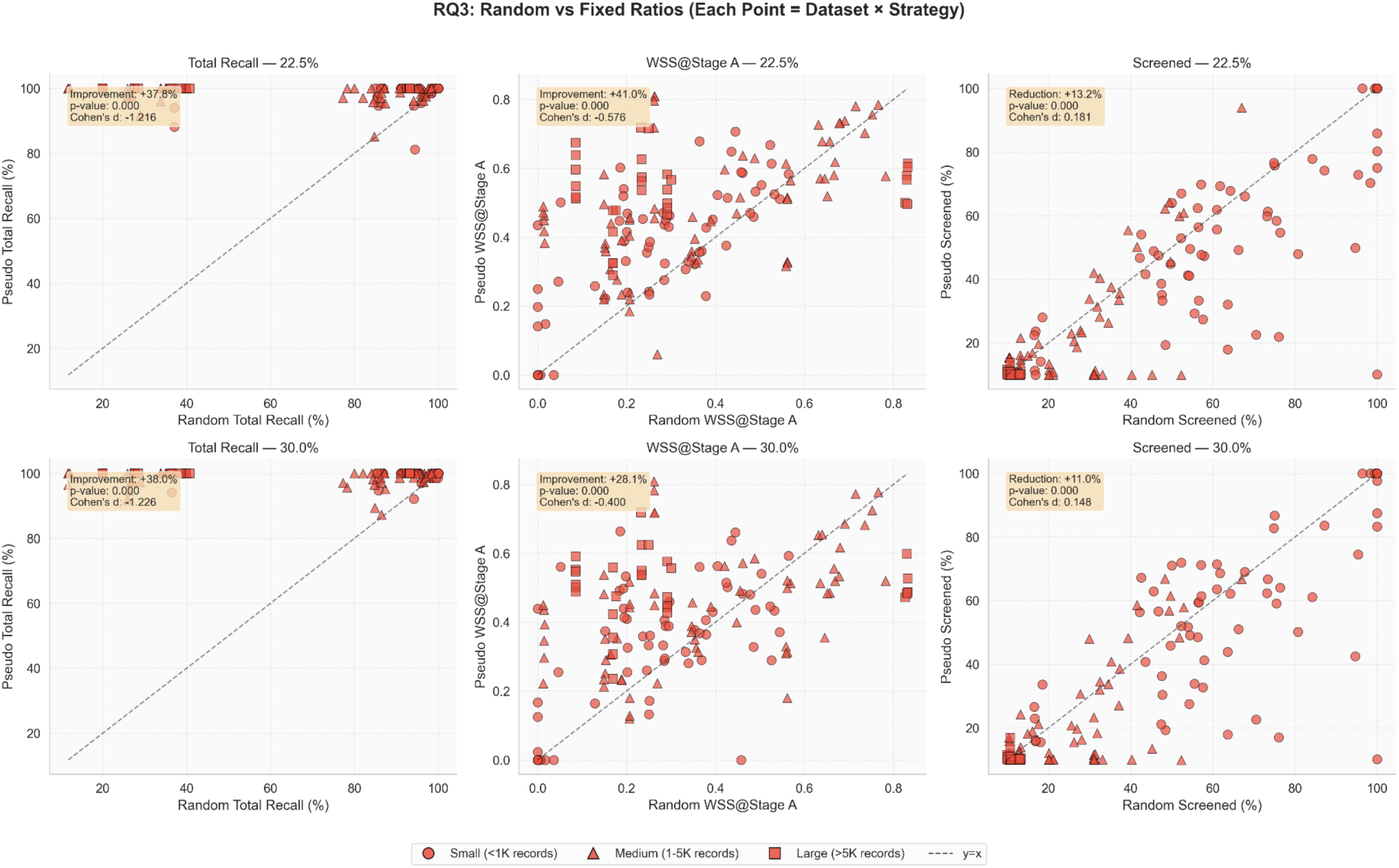
Pairwise scatter plots of outcomes for random versus pseudo-labelled initialisation (Universal 22.5%, top panel; Fixed 30.0%, bottom panel), across all 168 strategy-dataset pairs (28 datasets × 6 strategies). Left column: Total Recall (%); Centre column: WSS@Stage A; Right column: Screened (%). For Total Recall and WSS@Stage A (left and centre columns), points above the *y = x* diagonal indicate improvement under pseudo-labelling, since higher values are better. For Screened (%) (right column), points below the *y = x* diagonal indicate improvement, since lower values are better. Improvement/reduction percentages and Cohen’s *d* are shown in each panel.

The total recall scatter plots (left column) reveal the most dramatic pattern: the overwhelming majority of strategy-dataset pairs shift from widely dispersed recall values under random initialisation (approximately 10-100%) to near-ceiling values under both pseudo-labelling conditions (clustered at 95-100%). The improvements are consistent across both ratio conditions (a relative 37.8% increase for Universal 22.5% and 38.0% for Fixed 30.0%, respectively), with large values of Cohen’s *d* of −1.216 and −1.226, respectively (negative sign because pseudo-labelling is plotted on the *y*-axis and higher values are better), confirming that the effects are both pervasive and substantial.

The WSS@Stage A scatter plots (centre column) show a similar but less concentrated improvement: most points lie above the diagonal, indicating higher work savings under pseudo-labelling, but with greater dispersion than total recall. A small minority of strategy-dataset pairs achieve lower WSS under pseudo-labelling. However, the reported WSS values for Random may not reflect the reality because Random achieves a poor success rate and suffers from high sensitivity to (the seed for) the initial randomisation.

Cohen’s *d* values (−0.576 at 22.5%, −0.400 at 30.0%) confirm a medium-to-small effect for WSS, compared to the large effect for recall.

The % Screened scatter plots (right column) show the greatest spread around the diagonal. While the majority of points fall below it (indicating fewer records screened under pseudo-labelling), a meaningful fraction falls above it. Again, recall that pseudo-labelling achieves almost perfect success rates and close-to-perfect recalls and there is no practical need to enter the F stage (for finding more positive samples as SAFE suggests), while using Random we must enter Stage F and continue screening in ∼60% of the scenarios. In other words, pseudo-labelling makes SAFE a much more practical stopping criterion with overall less screening workload (at least on average). Cohen’s *d* is small (0.181 at 22.5%, 0.148 at 30.0%), indicating that the screening burden reduction, while statistically significant and practically relevant at the aggregate level, is less uniform across individual dataset-strategy combinations than the recall gains are.

#### 5.3.3. Strategy-Level Outcomes

Table 7 disaggregates the overall outcomes by query strategy, reporting success rate and WSS@Stage A for each of the six strategies under all four initialisation conditions.

**Figure 5.**
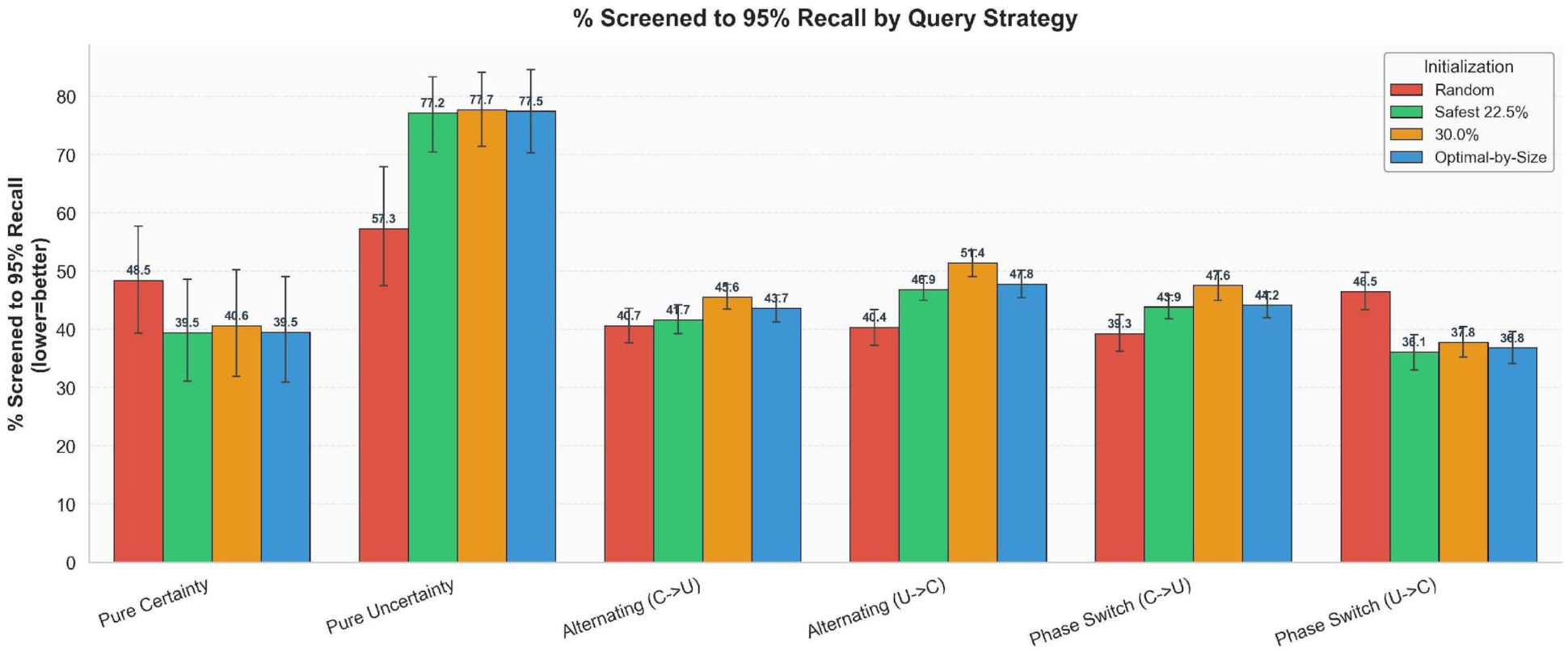
% Screened to 95% Recall by query strategy and initialisation condition. Lower values indicate faster convergence to 95% recall. Error bars = ±1 SD. Strategies are ordered by their convergence profile under pseudo-labelling; Pure Uncertainty shows the most severe worsening, while Phase Switch U→C and Pure Certainty show the greatest improvement.

**Figure 6.**
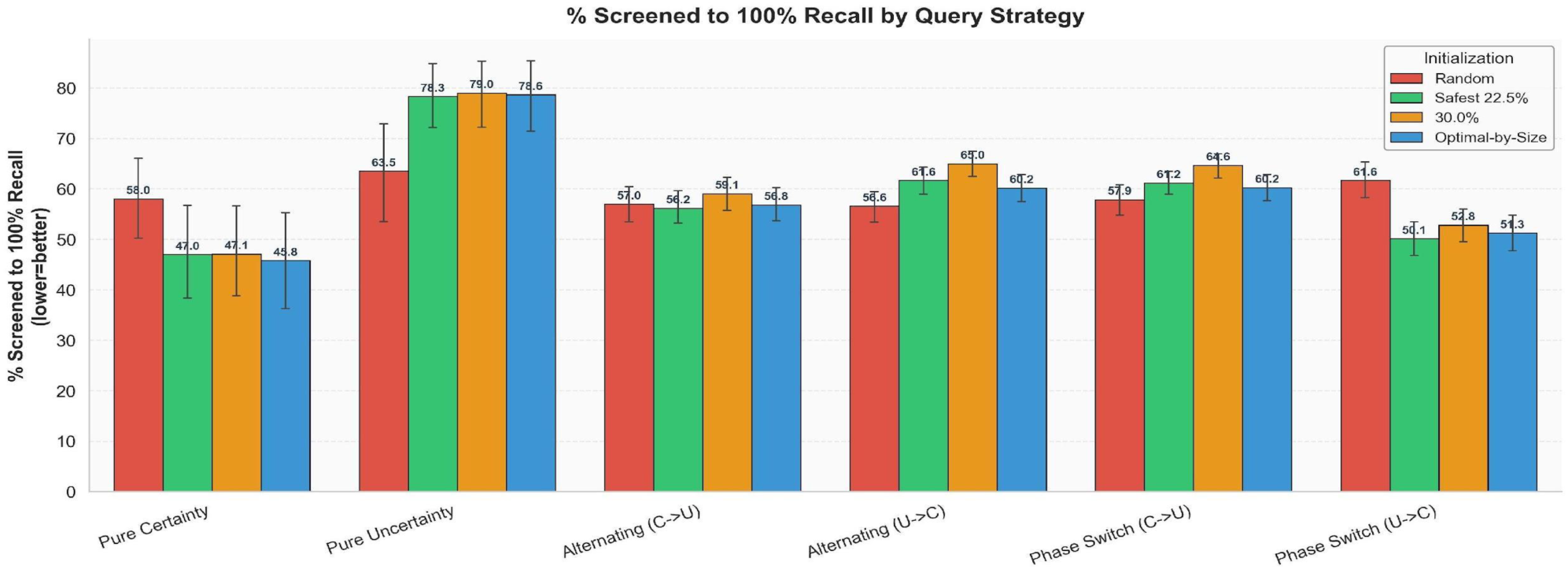
% Screened to 100% Recall by query strategy and initialisation condition. Lower values indicate that fewer records must be reviewed before all relevant documents are found. Error bars = ±1 SD.

**Figure 7.**
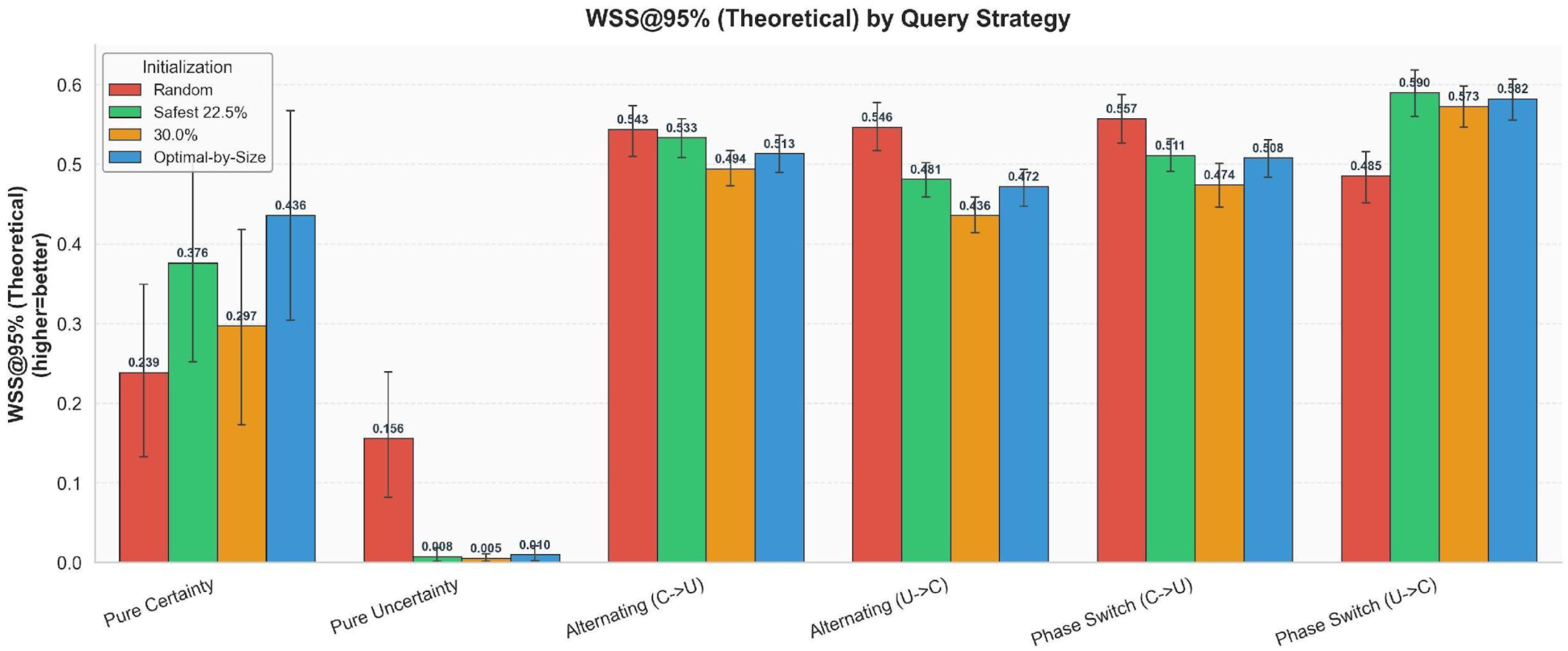
WSS@95% (Theoretical) by query strategy and initialisation condition. Higher values indicate greater work savings. Error bars = ±1 SD. The near-zero WSS values for Pure Uncertainty across all pseudo-labelling conditions indicate that almost no work is saved, i.e., the active learner reviews the corpus nearly exhaustively before reaching 95% recall.

Pseudo-labelled initialisation substantially improves success rates across every query strategy. Under random initialisation, success rates range from 35.7% (Pure Certainty, Pure Uncertainty) to 50.0% (Phase Switch U→C). Under Universal 22.5%, success rates rise to 89.3-100%, with both Alternating strategies reaching 100% while Phase Switch C→U lags at 89.3%. Under Fixed 30.0%, Pure Uncertainty, Alternating U→C, and Phase Switch U→C all reach 100%, while Pure Certainty drops to 85.7%. Optimal-by-Size achieves the broadest coverage: four of the six strategies reach 100% success (Alternating U→C, Alternating C→U, Pure Certainty, and both Alternating variants), and the remaining two (Phase Switch C→U and Phase Switch U→C) reach 96.4%.

WSS@Stage A patterns across strategies reveal a notable inversion relative to the reliability rankings established in RQ2. In RQ2, Pure Uncertainty ranked first on success rate when averaged across all 14 ratios. Here, at fixed best ratios, Pure Certainty and Phase Switch strategies achieve the highest WSS under pseudo-labelling (Pure Certainty: 0.471, Phase Switch U→C: 0.474, Phase Switch C→U: 0.445-0.446), while Pure Uncertainty achieves the lowest WSS (0.367-0.371), at least 10pp below the exploitation-oriented strategies. This reflects the mechanism discussed in RQ2: Pure Uncertainty’s boundary-exploration querying identifies relevant documents reliably but tends to review more records overall before triggering the stopping criterion. The Alternating strategies occupy an intermediate position (WSS 0.399-0.407), offering a balance between the high reliability of exploration-based querying and the work savings of exploitation-based strategies.

An important exception applies to Phase Switch C→U under Universal 22.5%, which achieves only an 89.3% success rate, the lowest among all strategy-initialisation combinations in the pseudo-labelling conditions, despite having WSS (0.445) among the higher values. This reflects the structural interaction between certainty-first querying and pseudo-labelled initialisation: starting with *exploitation*-focused sampling exhausts the high-confidence region early, potentially triggering the stopping criterion before all relevant documents are found in some datasets. Phase Switch C→U recovers to 96.4% success under the Fixed 30.0% and Optimal-by-Size conditions, suggesting that the higher ratio provides enough diversity in the initial training set to mitigate this early-exhaustion effect.

A secondary question concerns whether C-start and U-start strategies respond systematically differently to ratio choice. A formal strategy × ratio interaction test on the binary success outcome confirms that strategies do respond differently to ratio (logistic regression likelihood-ratio test: *χ*² = 13.30, *df* = 5, *p* = 0.021), but the pattern is not cleanly captured by the C-start versus U-start dichotomy. When the six strategies are grouped by their starting query mode, the start × ratio interaction is not statistically significant (*p* = 0.108 across all strategies; *p* = 0.060 for hybrid strategies only), and the direction of the marginal effect is opposite to a “U-start prefers smaller ratios” hypothesis. The driving observation is instead Pure Uncertainty’s unique behaviour: it reaches 100% strict success at 12.5%, the lowest ratio of any strategy, and maintains a near-flat success rate across the full ratio range (SD = 4.07pp across the 14 ratios, compared with 11.31-15.03pp for the other five strategies). All other strategies, regardless of the starting query mode, require ratios of 20.0-22.5% before reaching ≥95% strict success and exhibit substantially greater ratio sensitivity. On this measure, alternating strategies are not more reliable, i.e. less ratio-sensitive, than phase-switch strategies (Alternating U→C SD = 15.03 vs Phase Switch U→C SD = 14.93; Alternating C→U SD = 12.03 vs Phase Switch C→U SD = 11.31). The practical implication is that ratio choice matters more for some strategies than others, and that Pure Uncertainty’s robustness to ratio choice is a distinguishing property worth flagging for practitioners selecting a default configuration.

#### 5.3.4. Discussion: RQ3

##### 5.3.4.1. Why Stage S Gains Propagate to the Full Pipeline

The near-complete transfer of Stage S recall improvements to Stage A success rates reflects a structural alignment between the two stages. As demonstrated in RQ1, pseudo-labelled initialisation raises End-of-Stage-S classifier recall from approximately 14% to 99% across all datasets, meaning that the classifier entering Stage A has been trained on a set of examples that already identify the vast majority of relevant documents. This produces a classifier whose decision boundary at the start of Stage A is much closer to the true class boundary, leaving much less work for the active learning cycle to do.

This stands in sharp contrast to random initialisation, where the End-of-Stage-A classifier starts with a very limited number of positive examples (often only one or two positive samples), creating a shallow, poorly calibrated boundary. The active learning cycle must then compensate for this poor start by reviewing large numbers of documents before the boundary stabilises, resulting in low recall when the stopping criterion is met for a majority of datasets. The 56-58pp increase in success rate between random and pseudo-labelled conditions quantifies the magnitude of this cold-start problem and its resolution.

The modest but consistent reduction in screened proportion (relative reductions by −9.4% to −13.2%) reflects a secondary benefit: when the initial classifier is well-calibrated, the active learning cycle converges faster (in terms of reaching SAFE’s stopping criteria for Stage A), meaning fewer records need to be reviewed before the stopping criterion is triggered. However, this efficiency gain is substantially smaller and more variable across datasets than the recall gain, confirming that recall reliability is the primary mechanism through which pseudo-labelling improves the SAFE procedure, rather than a fundamental change in the number of documents reviewed per dataset.

##### 5.3.4.2. Why Universal 22.5% Outperforms Fixed 30.0% on Workload Savings

The finding that Universal 22.5% achieves a significantly higher WSS@Stage A (0.429) than Fixed 30.0% (0.390), despite identical success rates (95.8%), is consistent with the mechanism identified in RQ1 for Stage S performance. At 30.0%, a higher proportion of pseudo-labels are drawn from the middle of the LLM ranking, where scoring uncertainty is greatest and label noise is highest. This increased noise in the initial training set forces the active learning cycle to review more uncertain documents before the classifier boundary stabilises sufficiently to satisfy the SAFE stopping criteria. The additional label noise introduces more decision uncertainty near the boundary, which uncertainty-based strategies, in particular, must resolve through additional querying, extending the active learning phase and reducing overall WSS.

This result has practical consequences for ratio selection. Although both 22.5% and 30.0% achieve equivalent success rates at the aggregate level, choosing 22.5% over 30.0% produces meaningfully higher work savings (Δ = +0.039 WSS), equivalent to approximately additional 4 percent of the corpus saved per review. Across a systematic review corpus of, say, 5,000 records, this difference corresponds to approximately 200 fewer records reviewed, a practically important reduction in human screening effort.

##### 5.3.4.3. Strategy-Specific Mechanisms and the Reliability-Efficiency Trade-Off

The strategy-level WSS analysis reveals that pseudo-labelled initialisation amplifies rather than eliminates the reliability-efficiency trade-off observed in RQ2. Under random initialisation, all six strategies achieve low, broadly similar success rates (35.7-50.0%), with the Pure Certainty and Phase Switch strategies showing higher WSS due to their exploitation focus but lower recall. Under pseudo-labelling, success rates converge upward across all strategies (89.3-100% under Universal 22.5%). Still, the WSS advantage of exploitation-oriented strategies is preserved and enhanced: Pure Certainty reaches WSS = 0.471 and Phase Switch U→C reaches 0.474, while Pure Uncertainty remains at 0.371. The pseudo-labelled initialisation, therefore, provides a more reliable foundation for all strategies while allowing each strategy’s intrinsic mechanism to determine the efficiency of the subsequent active learning cycle.

The Phase Switch C→U strategy at Universal 22.5% constitutes an important exception: its 89.3% success rate, the lowest across all pseudo-labelling conditions, indicates that certainty-first querying under 22.5% initialisation creates a risk of premature stopping. The strategy’s recovery to 96.4% under Fixed 30.0% and Optimal-by-Size suggests that a larger, more diverse initial training set can mitigate this risk, as the wider coverage of pseudo-labels at higher ratios provides the End-of-Stage-A classifier with sufficient boundary examples, preventing early exhaustion of certainty-based querying. Practitioners selecting Phase Switch C→U should therefore prefer Fixed 30.0% or the size-adaptive ratio over Universal 22.5%.

##### 5.3.4.4. Practical Implications for the SAFE Procedure

In response to RQ3, LLM-based pseudo-labelling significantly enhances the SAFE procedure across all six query strategies and under both fixed- and size-adaptive-ratio conditions, which is achieved without any change to the active learning algorithm or the SAFE stopping criterion: pseudo-labelling operates entirely as an initialisation layer, requiring no modification to the core screening workflow.

For practitioners, the evidence supports a safe ratio range of approximately 22.5%–30.0% as the most practical recommendation. Within this range, strict success rates remain at or above 94.6%, relaxed success rates at or above 97.6%, and total recall at or above 99.1% (Table 2), with all configurations resolving the cold-start failure that affects the original SAFE procedure. Within this range, lower ratios are preferable when work savings are the priority: Universal 22.5% achieves the highest WSS@Stage A (0.429) and is statistically significantly higher than Fixed 30.0% (Δ = +0.039, *p* < 0.001) despite identical strict success rates (95.8%). Higher ratios within the range provide a small additional reliability margin against near-miss failures, with relaxed success rising to 98.8% at 30.0% and 99.4% at 27.5%, which may be valuable for reviews where the cost of missing a relevant study is particularly high.

When the dataset size is known in advance, size-adaptive ratios (32.5% for small, 22.5% for medium, 15.0% for large) provide a marginal additional improvement in success rate (+2.4pp over Universal 22.5%) at a modest cost in WSS for small and large datasets, and should be preferred when the highest possible reliability is required. In all cases, pseudo-labelled initialisation eliminates the cold-start problem that prevents the original SAFE procedure from achieving reliable abstract screening across most benchmark datasets, without requiring any additional human annotation before screening begins.

##### 5.3.4.5. Stage S vs Stage A: Is Active Learning Still Necessary?

The Stage S vs Stage A comparison yields four interconnected conclusions that together define the proper role of LLM pseudo-labelling in the systematic review pipeline.

First, Stage S is a powerful but incomplete screening tool. Its ability to achieve 92.9%–100% success rates and near-identical recall to Stage A, while requiring zero human screening, demonstrates that LLM-based ranking alone provides substantial scientific value. However, the irreversibility of its errors, its incompatibility with human-verification requirements and the loss of chance of human learning during reviewing mean it cannot serve as a standalone replacement for active learning.

Second, the pseudo-labelling ratio is the primary lever governing system performance. At low ratios, Stage S outperforms Stage A because active learning begins without sufficient initialisation signal; at moderate-to-high ratios, Stage A recovers and matches or surpasses Stage S by leveraging richer pseudo-label priors.

This confirms the rationale developed under RQ2: the pseudo-labelling ratio should be treated as a foundational configuration decision rather than a secondary tuning parameter.

Third, WSS convergence between Stage S and Stage A is mechanistically informative rather than coincidental. Stage S defers its efficiency cost to broader positive predictions; Stage A pays its cost upfront through human querying but recovers precision over iterations. These dynamics equalise net WSS across most ratios, with Stage A regaining the advantage at higher ratios as the active learning budget remains sufficient to correct accumulated pseudo-positive noise.

Fourth, the value of Stage A is methodological and epistemic, not metric-driven. Because Stage S achieves near-identical recall in many configurations, the genuine argument for retaining Stage A is that it provides verified human judgments that satisfy evidence synthesis standards and progressively corrects LLM errors that would otherwise be undetectable. These properties are not reflected in the simulation metrics in prior studies.

### 5.4. RQ4: Does Pseudo-Labelling Allow Active Learning to Converge Faster?

RQ4 examines whether pseudo-labelled initialisation accelerates active learning convergence, measured as the proportion of the corpus that must be screened before the stated recall threshold is reached: 95% recall (% Screened to 95% Recall), 100% recall (% Screened to 100% Recall), and the theoretical Work Saved over Sampling at 95% recall (WSS@95% Theoretical). Unlike RQ3, which assesses end-to-end screening outcomes at the point of SAFE stopping, RQ4 measures how rapidly relevant documents accumulate as documents are screened in order, tracking the point at which the cumulative recall threshold is crossed, which is a dominant approach adopted by prior simulation studies. This is an intrinsic property of the query ordering produced by the active learning strategy, evaluated across the full corpus independently of any stopping criterion. It therefore provides a theoretically grounded measure of retrieval efficiency, though it is worth noting that in practice, no stopping criterion can reliably identify the moment at which all positives have been found; the practical value of this analysis is in understanding how the query strategy distributes the discovery of relevant documents across the screening sequence. Again, three pseudo-labelling conditions are compared against random initialisation: Universal 22.5%, Fixed 30.0%, and Optimal-by-Size.

#### 5.4.1. Overall Convergence

Table 8 summarises the three convergence metrics for each initialisation condition, averaged across all strategies and datasets. Supplementary File 2 in the Appendix contains the results for all ratios. Across all three metrics and all pseudo-labelling conditions, the overall effect of pseudo-labelled initialisation on convergence speed is negligible or absent in the traditional context of simulation studies akin to prior studies.

**Table 8.**
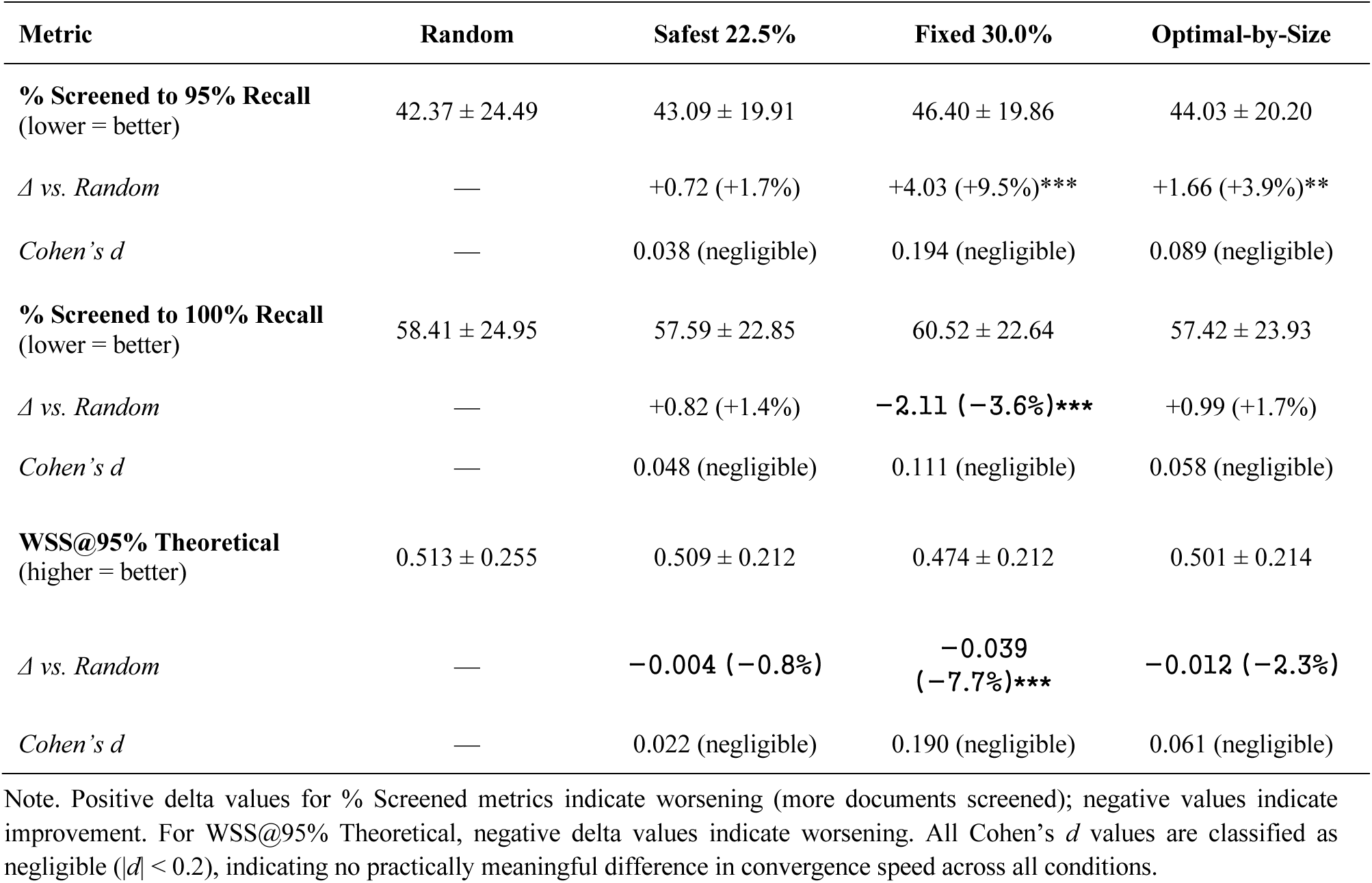
Convergence metrics under random initialisation versus three pseudo-labelling conditions, averaged across all six query strategies (*n* = 952 matched quadruplets). % screened = mean percentage of corpus screened before the stated recall threshold is reached. Mean ± SD reported for each condition. Significance: *** *p* < 0.001; ** *p* < 0.01 (Wilcoxon signed-rank vs Random). Statistically significant worsening relative to random is noted with *** *p* < 0.001.

The Universal 22.5% ratio shows no statistically significant difference from random initialisation on any convergence metric, with large *p* values far larger than 0.1 and negligible effective sizes, showing that the ratio that optimises screening reliability (Universal 22.5%) has no discernible effect on convergence speed when averaged across all strategies.

The Fixed 30.0% ratio, by contrast, produces statistically significant *worsening* of all three convergence metrics relative to random initialisation: more documents must be screened before reaching both 95% recall (Δ = −4.03pp, *p* < 0.001) and 100% recall (Δ = −2.11pp, *p* < 0.001), and WSS@95% Theoretical decreases by 0.039 (*p* < 0.001). However, all effect sizes remain negligible (*d* ≤ 0.194), indicating that these are statistically detectable but practically small differences, attributable primarily to increased label noise introduced by pseudo-labels drawn from the mid-ranked region of the LLM output.

The Optimal-by-Size condition yields mixed results: convergence to 95% recall worsens marginally (Δ = −1.66pp, *p* = 0.001), while convergence to 100% recall improves slightly (Δ = +0.99pp, *p* = 0.07, insignificant), and WSS@95% Theoretical shows a negligible reduction (Δ = −0.012, *p* = 0.06, insignificant). All effect sizes are negligible (*d* ≤ 0.089). These results confirm that size-adaptive ratio assignment does not systematically alter convergence speed in either direction, offering neither a convergence advantage nor a convergence cost relative to random initialisation.

#### 5.4.2. Strategy-Level Convergence

The aggregate neutrality of pseudo-labelled initialisation with respect to convergence may conceal sharp, mechanistically important divergences across query strategies. Figures 5-7 visualise the patterns for % Screened to 95% Recall, % Screened to 100% Recall, and WSS@95% Theoretical, respectively, by disaggregating the convergence metrics by strategy and initialisation condition.

Two strategies show clear improvement in convergence under pseudo-labelled initialisation. Pure Certainty reduces % Screened to 95% Recall from 48.47% (random) to 39.45% (Safest 22.5%), 40.63% (Fixed 30.0%), and 39.52% (Optimal-by-Size), representing improvements of approximately 8-9pp across all conditions. Correspondingly, % Screened to 100% Recall falls from 58.00% to 46.97-47.09%, and WSS@95% Theoretical increases from 0.239 to 0.376-0.436. Phase Switch U→C shows a similarly consistent pattern: % Screened to 95% Recall falls from 46.53% to 36.12-37.80%, WSS increases from 0.485 to 0.573-0.590. For these two strategies, pseudo-labelled initialisation provides a meaningful and consistent convergence benefit in addition to the reliability gains documented in RQ3.

Three strategies show modest-to-moderate worsening. Alternating C→U is for % Screened to 95% Recall near-neutral under Universal 22.5% (Δ = +1.01pp), but worsens under Fixed 30.0% (+4.92pp) and Optimal-by-Size (+3.02pp), suggesting that the additional label noise at higher ratios disrupts its exploitation–exploration balance. Alternating U→C worsens more clearly under all pseudo-labelling conditions (+6.51pp at Universal 22.5%, +11.08pp at Fixed 30.0%), consistent with the progressive shift from uncertainty- to certainty-based sampling being disrupted by an initialisation that has already imposed a structured boundary. Phase Switch C→U worsens by +4.6 to +8.3pp on % Screened to 95%, with commensurate reductions in WSS@95% Theoretical (from 0.557 to 0.474-0.511), though it showed strong reliability gains in RQ3.

Pure Uncertainty shows by far the most severe “deterioration” under pseudo-labelled initialisation. % screened to 95% Recall increases from 57.31% (random) to 77.20% (Uncertainty 22.5%), 77.69% (Fixed 30.0%), and 77.47% (Optimal-by-Size), representing a worsening of approximately 20pp under all pseudo-labelling conditions. % Screened to 100% Recall rises similarly, from 63.49% to approximately 78-79%. Most strikingly, WSS@95% Theoretical collapses from 0.156 (random) to 0.008 (Safest 22.5%), 0.005 (Fixed 30.0%), and 0.010 (Optimal-by-Size), approaching zero under all pseudo-labelling conditions.

The results corroborate with prior studies that claimed better convergence speed for active learning in simulation studies (Miwa et al., 2014). In our simulation studies, queries strategies based on or switching to certainty-based sampling “converge” faster with pseudo-labelling than random initialisation, but slower by strategies involving more or based on uncertainty-based sampling. However, it must be noticed that random initialisation fails to achieve the recall requirement on most tested reviews, and the success rate of certain-based sampling is still not satisfactory although being substantially improved over random initialisation (See Sect. 5.2.7.3 about the structural limitations of certainty-based sampling). On the contrary, success rates are substantially improved when using query strategies based on or focused on uncertainty-based sampling (Pure Uncertainty or Phase Switch C→U, respectively) or alternating between uncertainty-and certainty-based sampling (Alternating U→C or Alternating C→U) (See Sect. 5.2.7). So, if a different notion of “convergence” is taken, these query strategies can be said to “converge” faster with pseudo-labelling than random initialisation because (1) the SAFE procedure can “safely” stop after Stage A and (2) the percentages of samples screened at the end of Stage A for these query strategies with pseudo-labelling are either comparable to or smaller than random initialisation, as Figure 8 shows. This can also be seen in Figure 3, where Screened (%) is averaged across all query strategies. The subsequent subsections present discussions in more detail.

**Figure 8.**
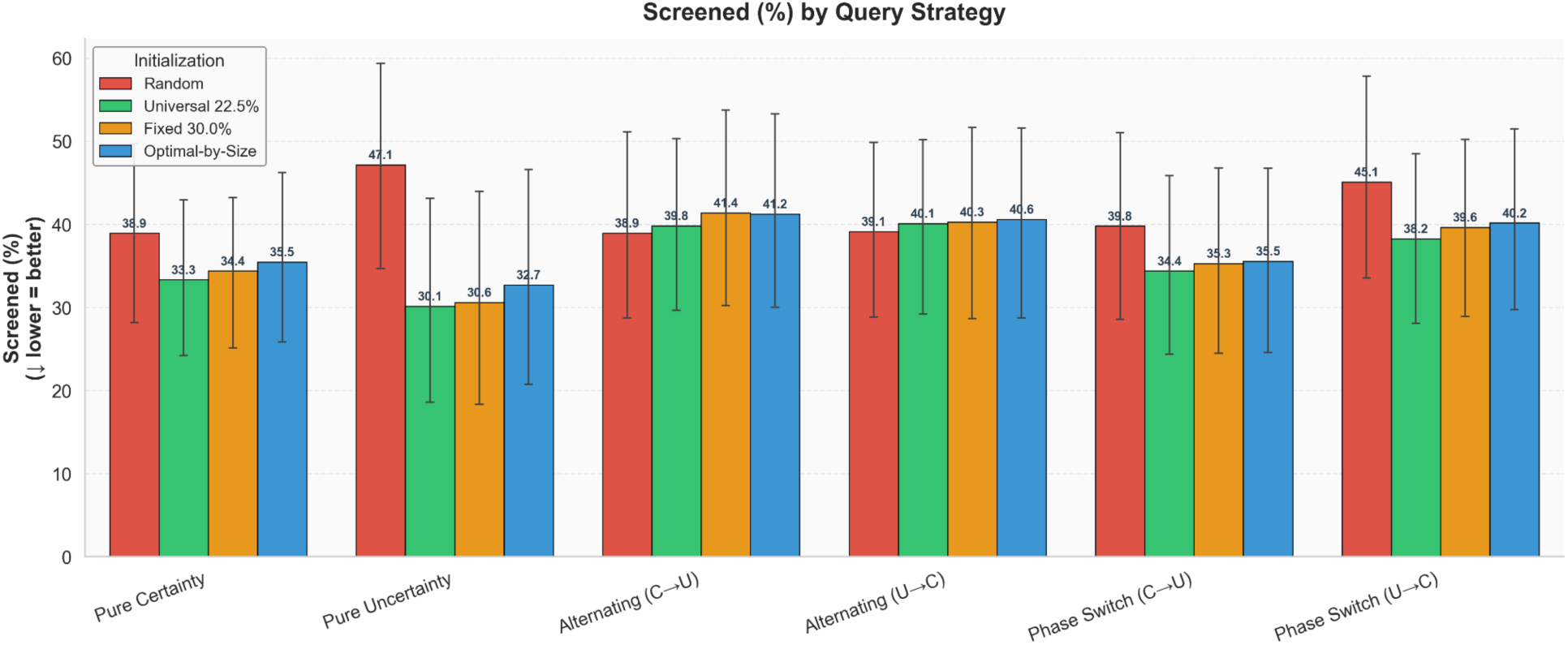
Percentage of corpus screened at the point SAFE stopping criteria were satisfied, by query strategy and initialisation condition, averaged across the datasets. Red bars represent random initialisation and bars in other colours represent pseudo-labelling with different ratios. Error bars represent ±1 SD. Lower values indicate that stopping was reached after less manual screening.

#### 5.4.3. Discussion: RQ4

##### 5.4.3.1. The Limits of Convergence as a Practical Measure

The overall finding that pseudo-labelled initialisation leaves convergence speed statistically unchanged (Safest 22.5%) or marginally worsened (Fixed 30.0%) at the aggregate level must be interpreted with an important caveat that applies to the entire RQ4 analysis: the convergence metrics used here are theoretical constructs, not operational outcomes. % Screened to 95% Recall and % Screened to 100% Recall measure the point in the screening sequence at which a fixed recall threshold is crossed. Computing these metrics requires knowing, in advance, the total number of relevant documents in the corpus. In real systematic review practice, this information is unavailable until screening is complete.

This is not a minor technical limitation but a fundamental constraint on the practical applicability of convergence-based evaluation. Any screening workflow that attempted to stop at the 95% convergence point would need to solve exactly the problem that makes systematic review screening labour-intensive in the first place: reliably identifying when all relevant documents have been found. The RQ4 metrics are therefore best understood as characterising the intrinsic retrieval trajectory of each query strategy under each initialisation condition, a theoretically informative lens on the ordering of relevant document discovery.

Rather than relying on an unobservable recall threshold, SAFE operationalises the stopping decision through practical criteria that the reviewer can evaluate in real time, such as consecutive runs of screened documents without a new positive and classifier confidence stabilisation, the two available criteria used in the current study that can be applied consistently. By comparing the high success rates in RQ3 with RQ4 results, we argue that the metric % Screened to Reach 95%/100% Recall and simulation studies have the risk of failing to reveal real convergence speed: while simulation studies do not show a clear advantage of in convergence speed in convergence speed, active learning can empirically “converge” much faster than simulation shows because in the SAFE procedure Stage A can stop much earlier and the Stage-A classifier can identify nearly all included studies only at the cost of introducing very limited additional screening workload. Overall, SAFE integrated with LLM-guided pseudo-labelling results in a statistically significant screening workload reduction (See the conclusion for RQ3)

##### 5.4.3.2. Why the Convergence Patterns Differ Across Query Strategies

Despite the theoretical limitations of the convergence framing, the strategy-level patterns in RQ4 are mechanistically informative and worth understanding, particularly the sharp divergence between certainty-based and uncertainty-based strategies.

Certainty-based strategies, specifically Pure Certainty and Phase Switch U→C, show improved convergence under pseudo-labelled initialisation because there is a natural alignment between what these strategies query and where the positives are located. A well-calibrated initial classifier assigns high relevance scores to likely positive documents; certainty-based sampling queries these documents first. The result is that positives are discovered early in the screening sequence, reducing the proportion of the corpus that must be reviewed before the recall threshold is crossed. For Phase Switch U→C, the initial uncertainty phase quickly resolves the narrow residual uncertainty around the LLM-defined boundary, enabling an earlier switch to certainty-based sampling and compounding the benefit.

Uncertainty-based strategies interact with a well-calibrated classifier in the opposite way. Under pseudo-labelled initialisation, the relevant documents are concentrated at the high-certainty end of the classifier’s ranking, rather than near the decision boundary. Uncertainty-based sampling, which by design queries the documents the classifier is most uncertain about, therefore targets the boundary region where positives are least likely to reside. The documents queried are predominantly borderline, near-irrelevant cases, and the positives remain unretrieved until the screening sequence reaches them. This produces the apparent deterioration in convergence observed for Pure Uncertainty, where % screened to 95% Recall rises from 57% to approximately 77%. However, it is important to emphasise that this is a property of the theoretical convergence metric, not a failure of the strategy in a practical screening context. Under SAFE, Pure Uncertainty achieves a 100% Stage A success rate at Fixed 30.0% (RQ3) by (Stage A) stopping at a much earlier point, demonstrating that criterion-based stopping has high chance to recover reliable recall even when the retrieval trajectory is less front-loaded, with relevant records found gradually instead of early on.

##### 5.4.3.3. Practical Implications for Strategy Selection

For practitioners using pseudo-labelled initialisation within the SAFE procedure, the convergence patterns in RQ4 offer secondary rather than primary guidance. The primary criterion for strategy selection should remain screening reliability as measured in RQ3. From that perspective, all strategies are substantially improved by pseudo-labelling, with success rates ranging from 89.3% to 100% depending on the strategy and ratio, compared with 35.7%–50.0% under random initialisation.

Where a practitioner additionally wishes to minimise the volume of documents reviewed during Stage A, the RQ4 results suggest that certainty-based and phase-switching strategies are preferable under pseudo-labelled initialisation, as they are more likely to retrieve positives early in the active learning cycle. Phase Switch U→C represents the strongest overall configuration, achieving a 96.4% success rate and the highest WSS@Stage A (0.474) in RQ3, with an 8-10 percentage-point improvement in convergence in RQ4. Pure Certainty with the Optimal-by-Size ratio offers a similar profile. The Alternating strategies remain viable where robustness is the primary concern.

In direct response to RQ4, pseudo-labelled initialisation does not systematically allow active learning to converge faster in the theoretical sense measured here. The more important observation, however, is that convergence speed in the sense of reaching an unobservable recall threshold is not a practically actionable measure for systematic review screening. The practical question is whether reliable recall can be achieved under a criterion-based stopping rule, and the evidence for RQ3 is unambiguous: pseudo-labelled initialisation raises SAFE success rates from below 40% to above 95% across all strategies and configurations. The convergence analysis in RQ4 complements this by showing how pseudo-labelling reshapes the retrieval trajectory, information that is useful for understanding the mechanics of each query strategy. Still, the criterion-based stopping results determine whether pseudo-labelled initialisation is suitable for deployment.

## 6. Limitations

Several limitations should be considered when interpreting these findings. First, all experiments are conducted on the CLEF eHealth 2019 benchmark, which comprises 28 systematic reviews in empirical medicine. Generalisation to reviews in other domains (e.g., social sciences, environmental health) or with substantially different prevalence rates cannot be assumed without further evaluation. Second, the active learning simulation uses the Naive Bayes classifier with TF-IDF feature extraction, the default ASReview configuration. Although this is the most widely benchmarked combination in the literature, the pseudo-labelling framework has not been evaluated against transformer-based classifiers or dense embedding representations, which may interact with pseudo-label noise in different ways. A companion study addressing this gap is currently in preparation. Third, only Stages S and A of the SAFE procedure are evaluated; the potential influence of pseudo-labelled initialisation on Stages F and E is not examined here and is left for future work. Fourth, the convergence metrics in RQ4 require knowledge of the total number of relevant documents in advance, which is unavailable in real screening practice; they should therefore be interpreted as characterising retrieval trajectories rather than providing actionable stopping guidance. Fifth, the LLM ensemble used for scoring (GPT-4o mini, Gemini 1.5 Flash, Claude 3 Haiku) was held fixed across all experiments; the framework’s sensitivity to LLM choice has not been systematically evaluated. Finally, the size-adaptive ratio recommendations are derived empirically from the 28-dataset benchmark and should be validated before being applied to reviews with substantially different corpus sizes or structural characteristics.

## 7. Conclusion

This study proposed LLM-based pseudo-labelling as a zero-annotation initialisation strategy for active learning-aided abstract screening and performed a comprehensive evaluation of the idea across 28 CLEF eHealth 2019 benchmark datasets and six query strategies, in total 4,032 experiments, in the context of the SAFE procedure for stopping active learning. The findings provide four clear takeaways for researchers and practitioners who adopt or extend active learning-based screening workflows.

First, pseudo-labelled initialisation is a reliable and effective replacement for random initialisation. The End-of-Stage-S classifier trained exclusively on pseudo-labels achieves 95% recall on 92.9%-100% of datasets across all tested ratios, compared to 7.5% under random initialisation, indicating it is a highly promising solution to the cold-start problem before any human annotation takes place.

Second, ratio selection matters and should be size-adaptive. Although a universal ratio of 22.5% is the safest fixed choice (95.8% Stage A success rate, highest WSS@Stage A among fixed ratios), but size-adaptive assignment, 32.5% for small datasets, 22.5% for medium, and 15.0% for large, raises the success rate to 98.2% overall and achieves 98.6%-100% success rates within each size category. Ratios above 22.5% introduce progressively more label noise from the mid-ranked region of the LLM output, reducing work savings without improving reliability.

Third, pseudo-labelled initialisation dramatically enhances the reliability of the end-to-end SAFE pipeline. Overall, Stage A success rates rise from 39.9% to 95.8%-98.2% (+56-58pp), total recall increases from 72.0% to 99.1%-99.3% (large effect, Cohen’s *d* = 0.86), and WSS@Stage A increases from 0.305 to 0.426- 0.429 (medium effect, Cohen’s *d* = 0.62). These gains require no modification to the active learning algorithm or the SAFE stopping criterion.

Fourth, convergence speed under pseudo-labelled initialisation varies by query strategy. Still, this finding should be interpreted with caution: the convergence metrics used in RQ4 are theoretical constructs that require knowing the total number of relevant documents in advance and being used only in simulation studies, which is unrealistic in real screening practice. This is precisely why structured stopping criteria such as SAFE are essential, as they provide an operational proxy for deciding when to stop without requiring knowledge of the true positive count. The RQ3 results, which evaluate performance under the SAFE stopping criterion directly, are therefore the more practically relevant basis for strategy selection: all six query strategies achieve substantially higher success rates under pseudo-labelled initialisation (89.3%-100%) than under random initialisation (35.7%-50.0%), and strategy choice should be driven primarily by this reliability evidence rather than by theoretical convergence trajectories.

Taken together, these results reframe initialisation, rather than query strategy or stopping criterion, as the principal determinant of whether SAFE-compatible active learning screening succeeds, and identify LLM-based pseudo-labelling as the mechanism that makes that initialisation reliable: it eliminates the need for prior human annotation, consistently achieves near-total recall at initialisation, raises screening reliability to near-ceiling levels, enables faster active learning convergence, and generalises reliably across datasets of varying sizes and prevalence rates requiring only an LLM relevance ranking of the candidate corpus and a single ratio decision before screening begins.

Future work will assess whether these benefits generalise across a broader range of feature representations and classifier architectures, whether observable properties of the candidate corpus can guide ratio selection, and whether the gains observed here hold under large-scale benchmarking on a more comprehensive set of medical research topics extending beyond the 28 CLEF eHealth 2019 reviews evaluated in the present study.

## Data Availability

All data produced in the present work are contained in the manuscript

## Notes

### Competing Interest Statement

The authors have declared no competing interest.

### Funding Statement

This study was funded by the Royal Society's International Exchange Scheme (IES/R1231175)

### Summary of Updates

This is a significantly modified version. The major revision was the integration of the idea of LLM-guided weak supervision with the SAFE procedure for reliable stopping of active learning for abstract screening. We evaluate the framework through 4,032 simulation experiments across 28 systematic reviews about clinical intervention and diagnostic technology assessment, spanning six query strategies and 14 pseudo-labelling ratios and answered four research questions about the reliability and recommended setups for applying LLM-guided weak supervision and SAFE procedure in real-world screening practice.

